# Defining multiple sclerosis subtypes using machine learning

**DOI:** 10.1101/19011080

**Authors:** Arman Eshaghi, Alexandra Young, Peter Wijertane, Ferran Prados, Douglas L. Arnold, Sridar Narayanan, Charles R. G. Guttmann, Frederik Barkhof, Daniel C Alexander, Alan J Thompson, Declan Chard, Olga Ciccarelli

## Abstract

Multiple sclerosis (MS) is subdivided into four phenotypes on the basis of medical history and clinical symptoms. These phenotypes are defined retrospectively and lack clear pathobiological underpinning. Since Magnetic Resonance Imaging (MRI) better reflects disease pathology than clinical symptoms, we aimed to explore MRI-driven subtypes of MS based on pathological changes visible on MRI using unsupervised machine learning. In separate train and external validation sets we looked at a total of 21,170 patient-years of data from 15 randomised controlled trials and three observational cohorts to explore MRI-driven subtypes and test whether these subtypes had differential clinical outcomes. We processed MRI data to obtain measures of brain volumes, lesion volumes, and normal appearing white matter T1/T2. We identified three MRI-driven subtypes who were similar in how they accumulated MRI abnormality. Based on the earliest abnormalities suggested by our model they were called: cortex-led, normal appearing white matter-led, and lesion-led subtypes. In the external validation datasets, the lesion-led subtype showed a faster disability progression and higher disease activity than the cortex-led subtype. In all datasets, MRI-driven subtypes were associated with disability progression (*β*_Subtype=_0.04, p=0.02; *β*_Stage=_-0.06, p<0.001), whilst clinical phenotypes and baseline disability were not. Only the lesion-led subtype showed a significant treatment response in three progressive multiple sclerosis randomised controlled trials (−66%, p=0.009) and in three relapsing remitting multiple sclerosis trials (−89%, p=0.04). Our results show that MRI-driven subtyping using machine learning can prospectively enrich clinical trials with patients who are most likely to respond to treatments.

## Introduction

More than 2.5 million people in the world live with multiple sclerosis (MS). Current thinking subdivides MS into four clinical phenotypes: clinically isolated syndrome (CIS), relapsing-remitting MS (RRMS), primary-progressive MS (PPMS) and secondary progressive MS (SPMS)^1^. These are categorised as relapsing (CIS and RRMS) or progressive (PPMS and SPMS) based on the current patient status and medical history. Two modifiers can be added to these categories: disease activity–as evidenced by relapses or new activity on magnetic resonance imaging (MRI)–and progression of disability^1^. Clinical phenotypes and their additional modifiers are routinely used in the clinical setting and in the selection of patients in clinical trials.

When examining the clinical, imaging, immunologic, or pathologic characteristics of the four MS phenotypes, the differences between the MS phenotypes are relative rather than absolute, because they are based on clinical descriptors, rather than on well-defined pathological mechanisms. A CIS phenotype may evolve into RRMS, and a RRMS phenotype may transition into SPMS.^1,2^ The precise timing of these transitions is challenging to ascertain, because they are often based on the *subjective* recollection of symptoms and their evolution. SPMS and PPMS share more similarities than differences in MRI features and pathogenic mechanisms.^3^ While most trials aim to recruit people with specific MS phenotypes, cohorts are still likely to contain people whose MS classification is not certain. Additionally, subgroup analyses of clinical trials have detected treatment effects for specific subgroups within phenotypes which are not seen when all patients were analyzed together^4,5^. The lack of well-defined boundaries between MS phenotypes has also introduced a misalignment between regulatory bodies^6^. Overall, there is a need for a greater sophistication and clarity in how MS phenotypes are defined.

In defining the phenotypes of MS more *objective* indicators of a patient’s biological status are urgently needed. MRI features are closer to the biology of MS than clinical symptoms, and better reflect the MS pathogenic mechanisms than purely clinical descriptions^7^. Therefore, we aimed to create a new framework, which defines the subtypes of MS based on the pathological changes visible on MRI scans, rather than clinical symptoms and course of disability over time. This objective subtyping framework will complement or modify the purely clinical course descriptors and represent an essential step towards personalised medicine, because it will lead to the use of therapies to target subpopulations who share the same pathogenic mechanisms of the disease and are most likely to benefit from these therapies^8^.

To explore whether it is possible to define MS subtypes on the basis of MRI abnormalities, we analysed a large number of MRI scans in patients with RRMS, SPMS and PPMS, using an unsupervised machine learning algorithm, called Subtype and Staging Inference (SuStaIn)^9^. Temporal change is a key barrier to identifying distinct subtypes of progressive diseases as “appearance of abnormality”, according to MRI, changes substantially over time; simply grouping individuals with similar data separates identical phenotypes at different time points. What makes SuStaIn different from all other unsupervised clustering algorithms is that it disentangles *temporal* change from *phenotypic* difference. SuStaIn identifies a set of subtypes each defined by a trajectory of change in various variables on a common time axis. SuStaIn extends the “event-based” models, previously used to identify single trajectories of change in MS^10^ and other neurodegenerative disorders^11,12^. After training, SuStaIn determines how closely information from a given patient matches ‘learned’ data-driven subtypes, and what stage the given patient has reached at a particular time. The ability to cluster patients using cross-sectional data makes SuStaIn a strong candidate for patient selection in clinical trials.

Here, we used SuStaIn to define a model that optimally explained the baseline MRI heterogeneity in 12 randomised controlled trials (RCTs) and two observational cohorts and identify subtypes. We then tested the trained model in three RCTs and one observational cohort, which were unseen (independent) datasets, thereby confirming its external generalisability. We aimed to: (1) to define MS *subtypes* on the basis of MRI-derived patterns, and *stage* a person’s progress within a subtype based on the trajectory of MRI changes; (2) Determine whether these MRI-driven subtypes–defined at *baseline*–and the standard clinical phenotypes predicted disease progression overt time; (3) Test whether there are differences in treatment response between these newly defined subtypes.

## Results

We present our results on training, internal and external validation of SuStaIn in the following sections (**Figure 1**):

**Figure 1.**
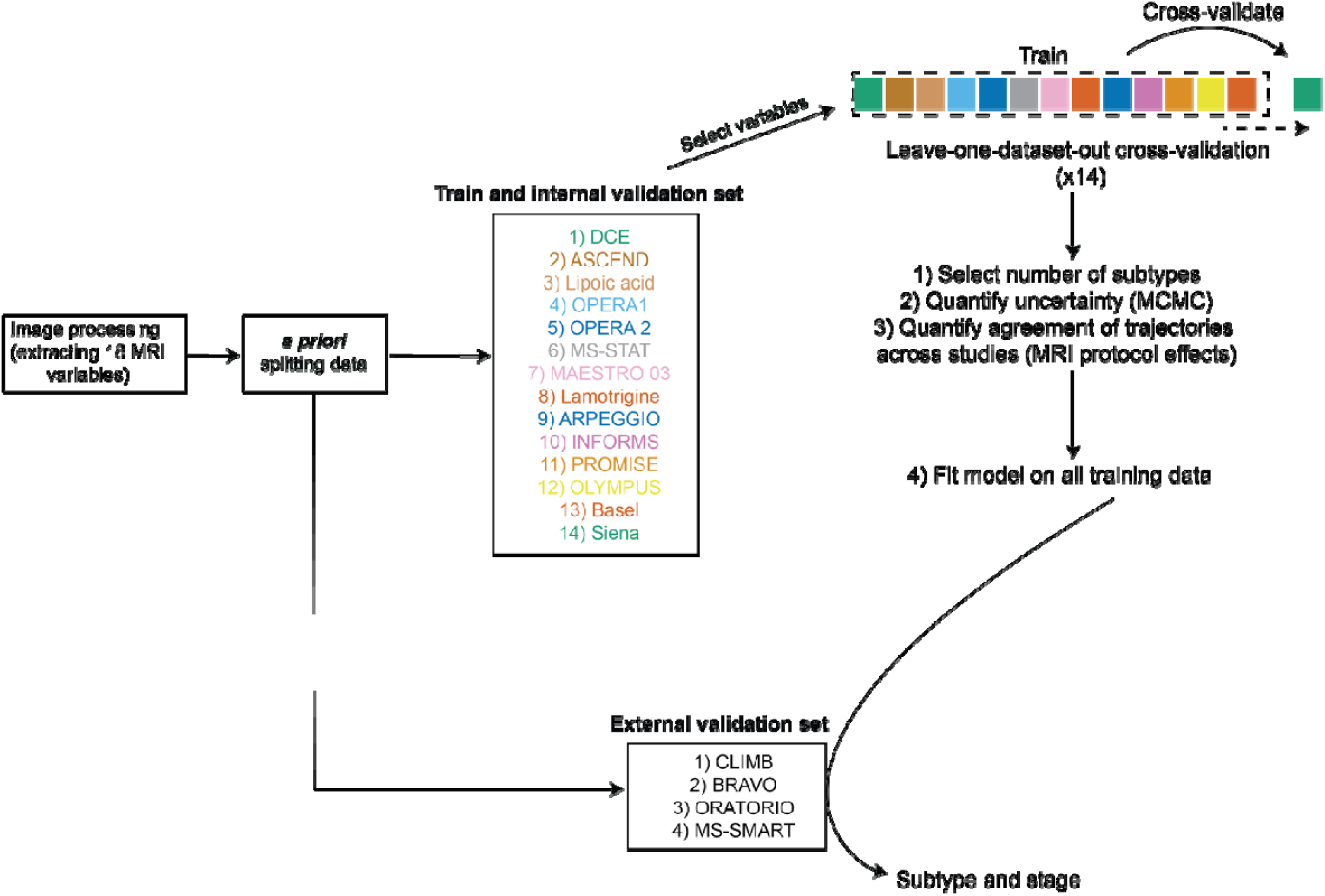
Model development. This figure shows that all “raw” data from different data sets underwent a unique image processing pipeline to extract MRI variables of lobar grey matter volume, visible white matter lesion from FLAIR, and T1/T2 ratio. We used healthy volunteers (UK Biobank and the Human Connectome Project) to adjust MRI measures nuisance variables (see main text), calculate Z-scores, and select MRI variables. *A priori* we split our patient datasets into two separate datasets: 14 datasets in the “train and internal validation” set, and three datasets in the external validation set: CLIMB (an observational study), BRAVO (a phase 3 RRMS trial), ORATORIO (a phase 3 PPMS trial), and MS-SMART (a phase 2 SPMS trial). Acronyms: MCMC, Markov Chain Monte Carlo; RRMS, relapsing remitting multiple sclerosis; PPMS, primary progressive multiple sclerosis; MRI, magnetic resonance imaging.

1. defining *a priori* internal and external validation datasets,
2. processing MRI data and selecting MRI variables,
3. training SuStaIn and internal validation on baseline MRI,
4. externally validating SuStaIn on independent, unseen datasets.

In this manuscript, we use “phenotype” when referring to standard clinical phenotypes, and “subtype” when referring to MRI-driven subtypes.

### Defining, *a priori*, training, internal, and external validation datasets

We retrospectively analysed data sets from 8,968 people with MS who had a total of 32,602 MRI visits from 18 datasets, which were 15 double-blind randomised controlled trials (RCTs) and three observational cohorts (**Table 1** and **Figure 1**). We split *a priori* these datasets into 6,322 patients (2,884 with RRMS, 1,837 with SPMS, and 1,601 with PPMS) in the train and internal validation set, which was used for model training and cross-validation, and 2,646 patients (1,512 RRMS, 711 PPMS and 423 SPMS) in the external validation set, which was used for model testing. **Table 1** shows included participants from these RCTs and observational cohorts. When we compared the train and internal validation set with the external validation set, patients in the external validation set were younger (average difference of 3.1 years, p<0.001), had shorter disease duration (average difference of 6 months, p=0.001), and were less disabled (0.5 difference in Expanded Disability Status Scale (EDSS), p<0.001) than those in the train and internal validation set.

**Table 1.** Collated datasets.

| **Study name** | **Population** | **Design** | **Participants**<br>**with eligible**<br>**MRI** | **Visits**<br>**with MRI** | **Published**<br>**protocol**<br>**citation**<br>**number** |
| --- | --- | --- | --- | --- | --- |
| **Datasets to develop norms** |  |  |  |  |  |
| UK Biobank* | Healthy<br>volunteers | Observational | 13,823 | 13,823 | <sup>32</sup> |
| Human<br>Connectome<br>Project* | Healthy<br>volunteers | Observational | 1,105 | 1,105 | <sup>33</sup> |
| **MS datasets in the train and internal validation set\*\*** |  |  |  |  |  |
| Siena | Mixed | Observational | 149 | 595 | <sup>38</sup> |
| Basel | Mixed | Observational | 81 | 239 | <sup>35</sup> |
| DEFINE-CONFIRM,<br>ENDORSE | RRMS | RCT<br>(phase 3) | 1,071 | 5,208 | <sup>29</sup> |
| OPERA 1 | RRMS | RCT<br>(phase 3) | 801 | 3,025 | <sup>30</sup> |
| OPERA 2 | RRMS | RCT<br>(phase 3) | 824 | 3,044 | <sup>30</sup> |
| ASCEND | SPMS | RCT<br>(phase 3) | 1,002 | 5,095 | <sup>46</sup> |
| Lipoic acid | SPMS | RCT<br>(phase 2) | 41 | 111 | <sup>26</sup> |
| MS-STAT | SPMS | RCT (phase 2) | 131 | 373 | <sup>24</sup> |
| MAESTRO 3 | SPMS | RCT<br>(phase 3) | 539 | 1,753 | <sup>27</sup> |
| Lamotrigine | SPMS | RCT<br>(phase 2) | 97 | 251 | <sup>25</sup> |
| ARPEGGIO | PPMS | RCT<br>(phase 2) | 409 | 946 | <sup>40</sup> |
| INFORMS | PPMS | RCT<br>(phase 3) | 323 | 758 | <sup>23</sup> |
| PROMISE | PPMS | RCT<br>(phase 3) | 458 | 740 | <sup>22</sup> |
| OLYMPUS | PPMS | RCT<br>(phase 2/3) | 396 | 1,630 | <sup>5</sup> |
| **MS datasets in the external validation set, for model testing\*\*** |  |  |  |  |  |
| CLIMB | Mixed | Observational | 319 | 1,950 | <sup>36</sup> |
| ORATORIO | PPMS | RCT<br>(phase 3) | 701 | 2,724 | <sup>13</sup> |
| BRAVO | RRMS | RCT<br>(phase 3) | 1,203 | 3,009 | <sup>31</sup> |
| MS-SMART | SPMS | RCT<br>(phase 2) | 425 | 1,151 | <sup>28</sup> |

\* UK Biobank and Human Connectome Project are cross-sectional cohorts. All others are longitudinal.
\*\* We chose training and external validation sets *a priori*.
\*\*\* Refers to MAESTRO 1 study, which has a similar protocol to MAESTRO 3 (unpublished).
Abbreviations: RCT=double-blind randomised controlled trial; RRMS=relapsing-remitting multiple sclerosis; SPMS=secondary progressive multiple sclerosis; PPMS=primary progressive multiple sclerosis; PMID, PubMed Identifier.

### MRI Processing: calculating normal ageing and gender effects using datasets of healthy volunteers

We obtained 18 MRI variables which were volumes of grey matter lobes and deep grey matter, white matter lesion load, and normal-appearing WM T1/T2 for all patients and healthy volunteers.

To estimate and adjust for demographic variables and ageing effects in MRI, we used two data sets from 14,928 healthy volunteers which covered a wide range of age (23.5 to 70 years, 13,823 from the UK Biobank and 1,105 from the Human Connectome Project; 7,965 women and 6,963 men) (**Table 1**). The mean age was 28.9 years (standard deviation=3.62) for the Human Connectome Project and 54.9 years (standard deviation=7.49) for the UK Biobank. We used healthy controls’ datasets to calculate linear and non-linear effects of age, gender, and total intracranial volume and adjusted MRI variables in patients for these effects. Of the 18 adjusted MRI variables, 13 were associated with a moderate to large effect size when patients at baseline visits were compared with healthy controls, and, therefore, were selected and entered into SuStaIn (**Figure 2a)**. Selected variables were volumes of the occipital, parietal, temporal, limbic and frontal grey matter, and deep grey matter; total white matter lesion volume; T1/T2 ratio in the corpus callosum, frontal, temporal, parietal, cingulate bundle and cerebellar normal-appearing white matter (NAWM) regions.

**Figure 2.**
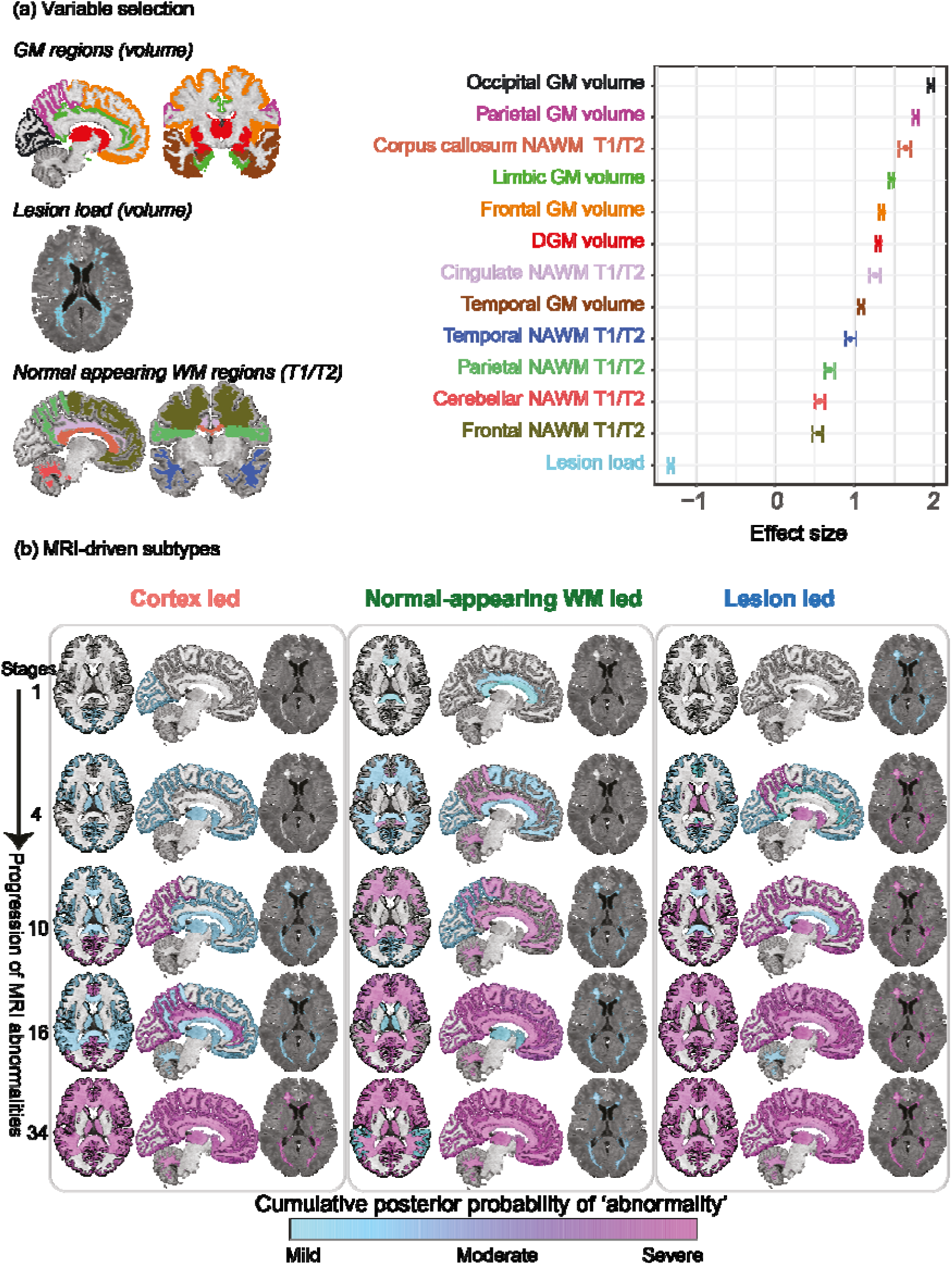
Variable selection and subtype trajectories. (a) We chose variables whose effect size was medium to large (Cohen’s d effect size greater than 0.5) when comparing all patients of the ‘train set’ with healthy volunteers. We have overlayed selected 13 variables on a T1-weighted MRI scan of a randomly chosen patient. We used the same colour coding to show selected variables on the brain MRI scan and the right plot. On the right plot, dots represent point estimates of the effect size and error bars represent the 95% confidence interval of the effect size. (b) This section shows the temporal sequence of MRI abnormalities in each of the three MRI-driven subtypes. The color shade ranges from blue to pink which represents the probability of ‘abnormality’ (it can be interpreted as the degree of ‘abnormality’) (mild, moderate or severe which approximates 1, 2 and, 3 sigma). The cortex-led subtype (left) showed cortical atrophy in the occipital, parietal and frontal cortex in the early stages of the sequences, and a reduction in T1/T2 ratio in the NAWM in the later stages. The normal-appearing white matter (NAWM)-led subtype (middle) showed a reduction in T1/T2 ratio of the cingulate bundle and corpus callosum in the earlier stages of the sequence, and deep grey matter and temporal grey matter atrophy in the later stages. The lesion-led subtype (right) shows early and extensive accumulation of lesions in the earlier stages of the sequence, and a reduction in the T1/T2 ratio in the NAWM in the later stages. The numbers on the left side represent SuStaIn stages. The minimum stage is 1 and the maximum stage is 39 (based on 13 variables that can show mild (sigma=1), moderate (sigma=2) and severe abnormality (sigma=3); 13×3 = 39). Acronyms: NAWM, normal-appearing white matter; SD, standard deviation; GM, grey matter; T1/T2, T1-T2 ratio.

### Training and internal validation: defining MRI-driven subtypes and the sequence of accumulation of MRI abnormality for each subtype

We used “leave-one-dataset-out” cross-validation on the baseline MRI data–which were acquired before administering experimental treatments–of the train and internal validation set to train our model and select the most optimal number of subtypes. The most optimal model in explaining heterogeneity in MRI variables had three subtypes (see Supplemental Results for details on model selection). Sequences of accumulation of MRI abnormalities in each of these three subtypes are shown in **Figure 2b** and **Supplementary Figure 1**. We called these three subtypes based on the variables that our model suggested to have earliest abnormality: the cortex-led, the normal-appearing white matter (NAWM)-led, and the lesion-led subtypes. The cortex-led subtype was characterised by an early cortical atrophy in the occipital, parietal, and frontal cortex, which was followed by atrophy in the other grey matter regions and T2 lesion accrual and, in the late stage, by a reduction in the T1/T2 ratio of the NAWM regions. The NAWM-led subtype was characterised by an early reduction in the T1/T2 ratio in the NAWM of the cingulate bundle and corpus callosum, which was then followed by a reduction in T1/T2 ratio in the cerebellar, temporal and parietal NAWM, atrophy of the occipital cortex, T2 lesion accrual, and, in the late stage, by atrophy of other cortical regions and deep grey matter. The lesion-led subtype was characterised by an early and extensive accrual of T2 lesions, which was followed by early severe deep grey matter atrophy, atrophy of the occipital, parietal and temporal cortex, and, in the late stage, by a reduction in NAWM T1/T2 ratio.

SuStaIn provided probabilistic assignment to three subtypes for each subject at baseline (upper section **Figure 3**). We assigned the subtype with the highest probability to each subject. **Table 2** shows the demographic, clinical and radiological characteristics of the three MRI-driven subtypes defined at baseline. The most frequent subtype was the cortex-led (43%). The lesion-led subtype had a slightly higher EDSS at baseline (median=4.5) compared to the other two subtypes (median EDSS for cortex-led 4.0, and NAWM-led=3.5, *p*<0.01). The lesion-led subtype had a longer disease duration (mean: 9.09 years) compared to the other two subtypes (6.27 years for the cortex-led and 5.56 years for the NAWM-led, *p*<0.01). When looking at MRI measures of each subtype, the lesion-led subtype showed the highest lesion load and the lowest cortical volume at baseline (p<0.001). Over time in the placebo arms, the lesion-led subtype had the fastest rate of cortical atrophy compared to the cortex-led and NAWM-led subtypes (p<0.001) and fastest rate of lesion accrual (p<0.001). The mean SuStaIn stage was slightly higher in the lesion-led subtype (average stage=17.9) compared to the cortex-led (average stage=15.9) and the NAWM-led subtype (average stage=13.7, p<0.01). There was a significant annual increase in stages within each subtype on RCT placebo arms (p<0.001 for all tests) and no changes between subtypes (**Table 2**).

**Table 2.**
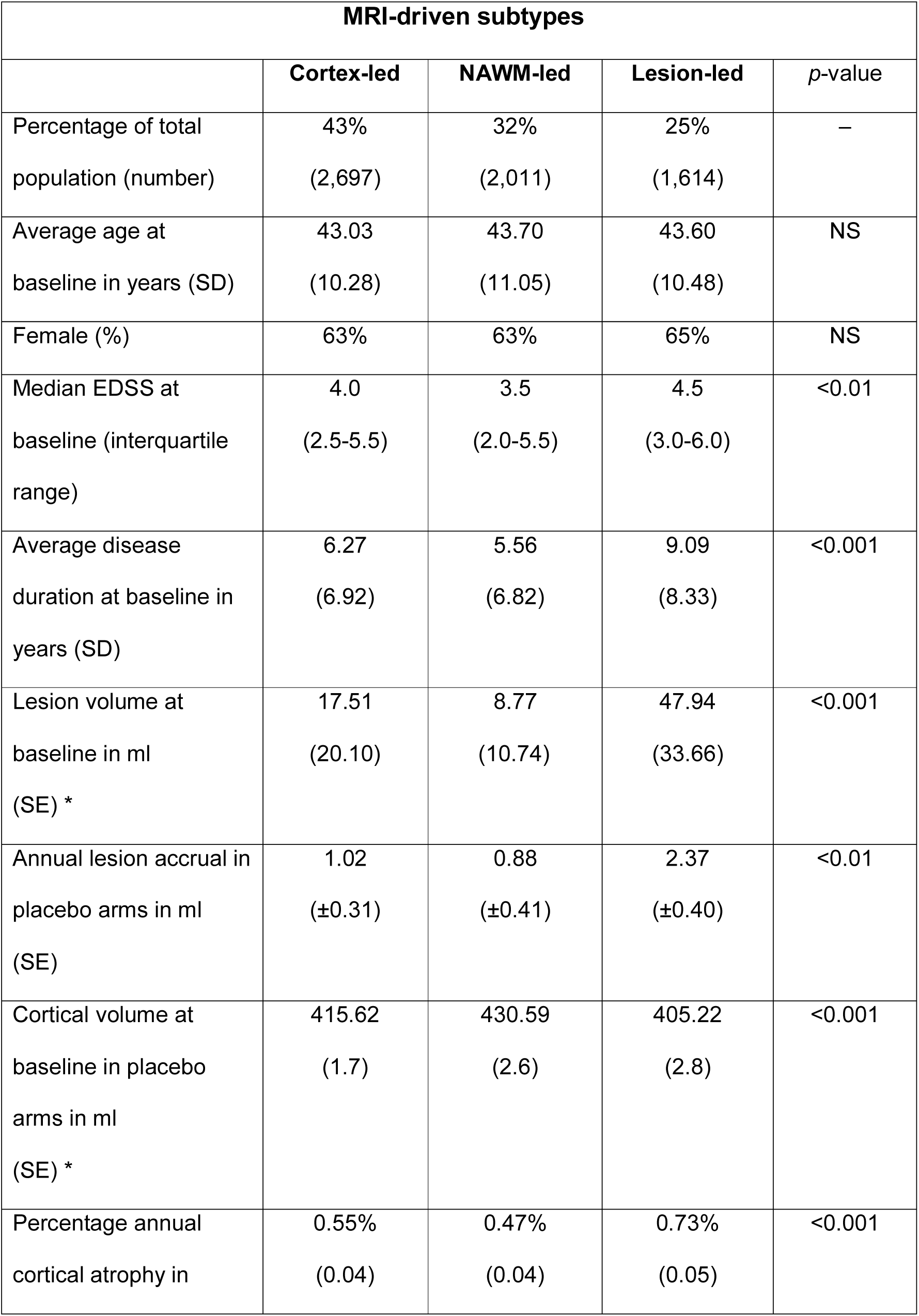
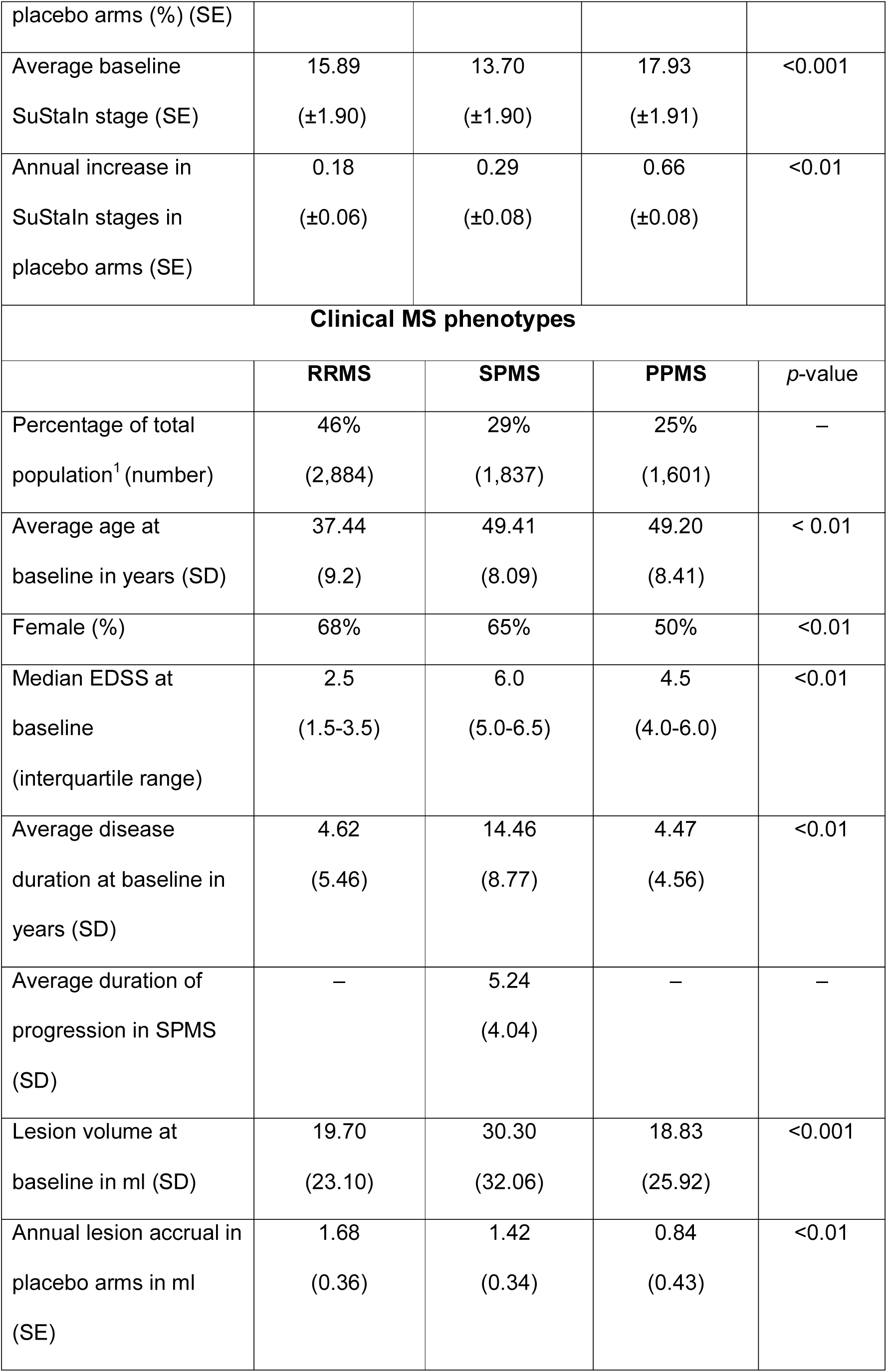
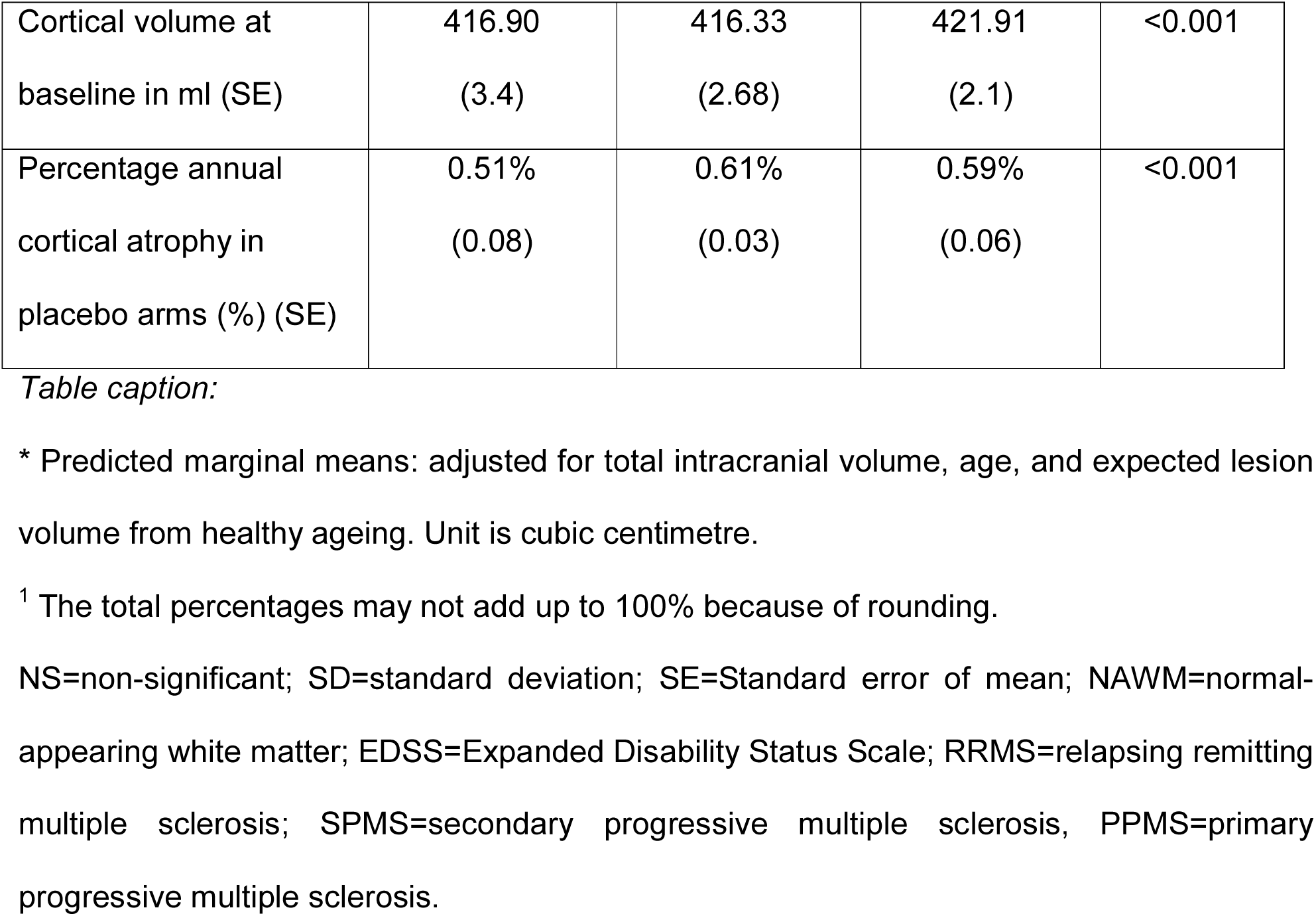
Demographic, clinical and radiological characteristics of the three MRI-driven subtypes and the clinical MS phenotypes in the train and internal validation set.

**Figure 3.**
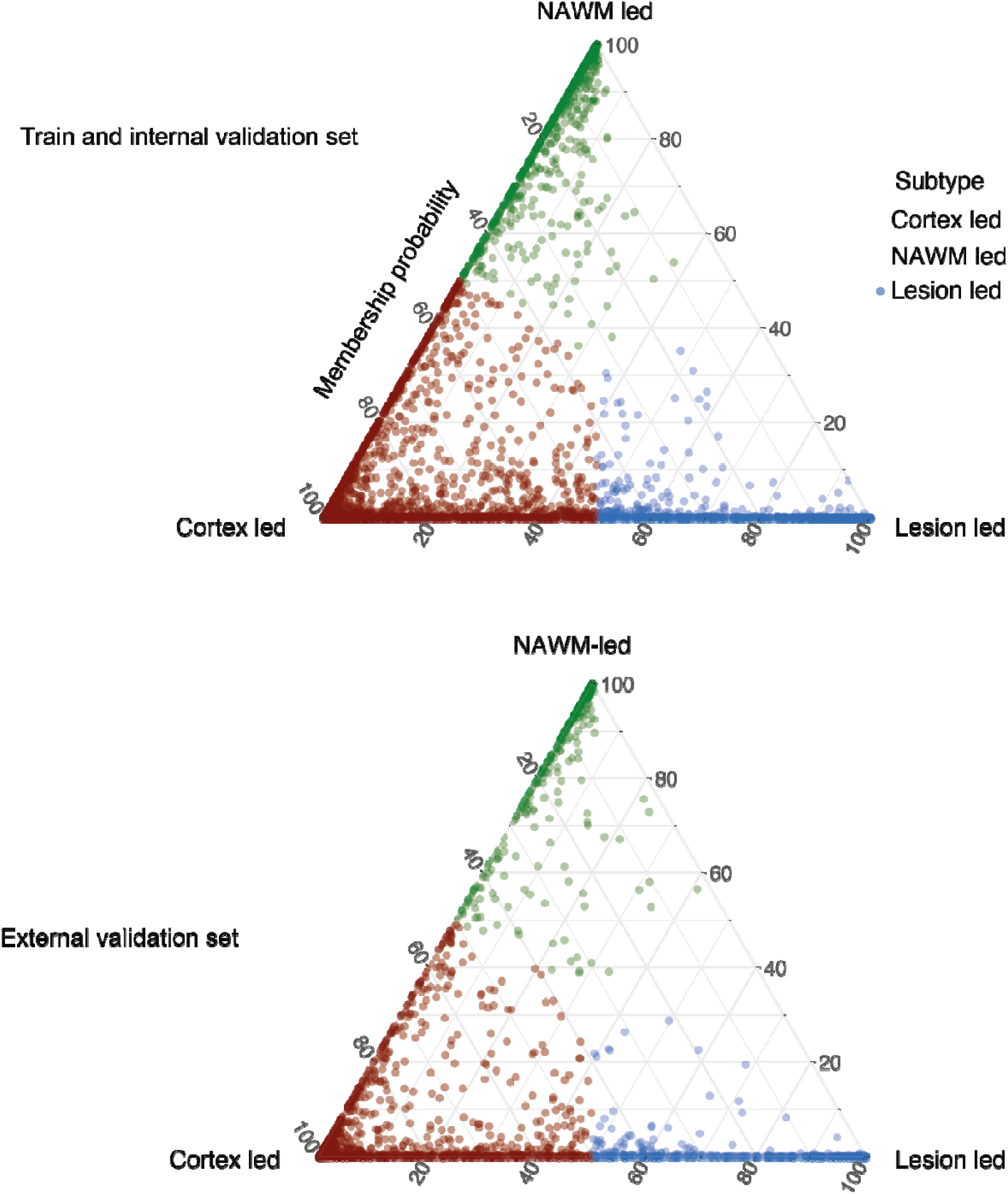
Subtype membership in the train, and external validation sets. MRI-driven subtypes in the train and cross-validation set (top) and the external validation set (bottom) are shown. Assignability or membership probability is shown as the distance from each vertex of the triangle; Each of vertices represent the point at which membership of a given subtype is at its maximum (100%). We assigned one subtype to each subject (shown in red, green and blue) based on the dominant membership.

When examining the standard MS phenotypes in the train and internal validation set (**Table 2**), most patients were RRMS. As expected, SPMS and PPMS were older and had higher EDSS than RRMS patients; additionally, SPMS patients had longer disease duration (time since diagnosis), higher T2 lesion load, and lower cortical volume than the other two phenotypes, and the fastest development of cortical atrophy (**Table 2**).

When exploring the distribution of MRI-driven subtypes across the standard MS phenotypes in the train and internal validation set (**Figure 3b**), the cortex-led subtype was the most common subtype in RRMS (46.9%) and the second most common subtype in both SPMS (39.7%) and PPMS (38.2%). The most common subtype in SPMS was the lesion-led subtype (41.7%), and in PPMS was the NAWM-led subtype (52.9%).

### “Dataset” and “centre” effects

We found that the subtypes were highly consistent across datasets in the train and internal validation set, despite different RCTs and trial protocols. In particular, the average measure of agreement (or Bhattacharyya coefficient) of the posterior distribution of the estimated sequences for each subtype across all cross-validation folds were as follows: 0.94 (standard deviation±0.03) for the cortex-led subtype, 0.94 (standard deviation ±0.02) for the NAWM-led subtype, and 0.96 (standard deviation ±0.02) for the lesion-led subtype, suggesting excellent agreement across trials. When we looked at the effects of centre inside each dataset on MRI-driven subtypes, the EDSS and MRI measures were significantly more strongly associated to “subtype” effect than the “centre effect” (see Supplemental Results).

### Generalisation ability of the model in the external validation set

When we tested the trained model the external validation set similarly to the train and internal validation set, the cortex-led was the most common subtype (42%), and the second most common subtype was the lesion-led subtype (37%), followed by the NAWM-led (20%) (see the lower section in **Figure 3**). The lesion-led subtype showed the longest disease duration, which was 1.4 years longer than cortex-led and 1.92 years longer than the NAWM-led subtype. The lesion-led subtype showed the largest lesion volume at baseline but lesion accrual over time was not significantly different across these subtypes. The cortex led and lesion led had lower cortical volumes at baseline than the NAWM-led, but this was not different between lesion-led and cortex-led. The cortex-led subtype had the fastest cortical atrophy over time. Similar to the train and internal validation set, the cortex-led subtype had the youngest age at baseline (1.7 years younger than the lesion-led and 2.75 younger that the NAWM-led subtype) and the lowest percentage of female patients (61%).The EDSS was similar across the three subtypes in the external validation set. **Table 3** shows the demographic, clinical and radiological characteristics of these subtypes.

**Table 3.**
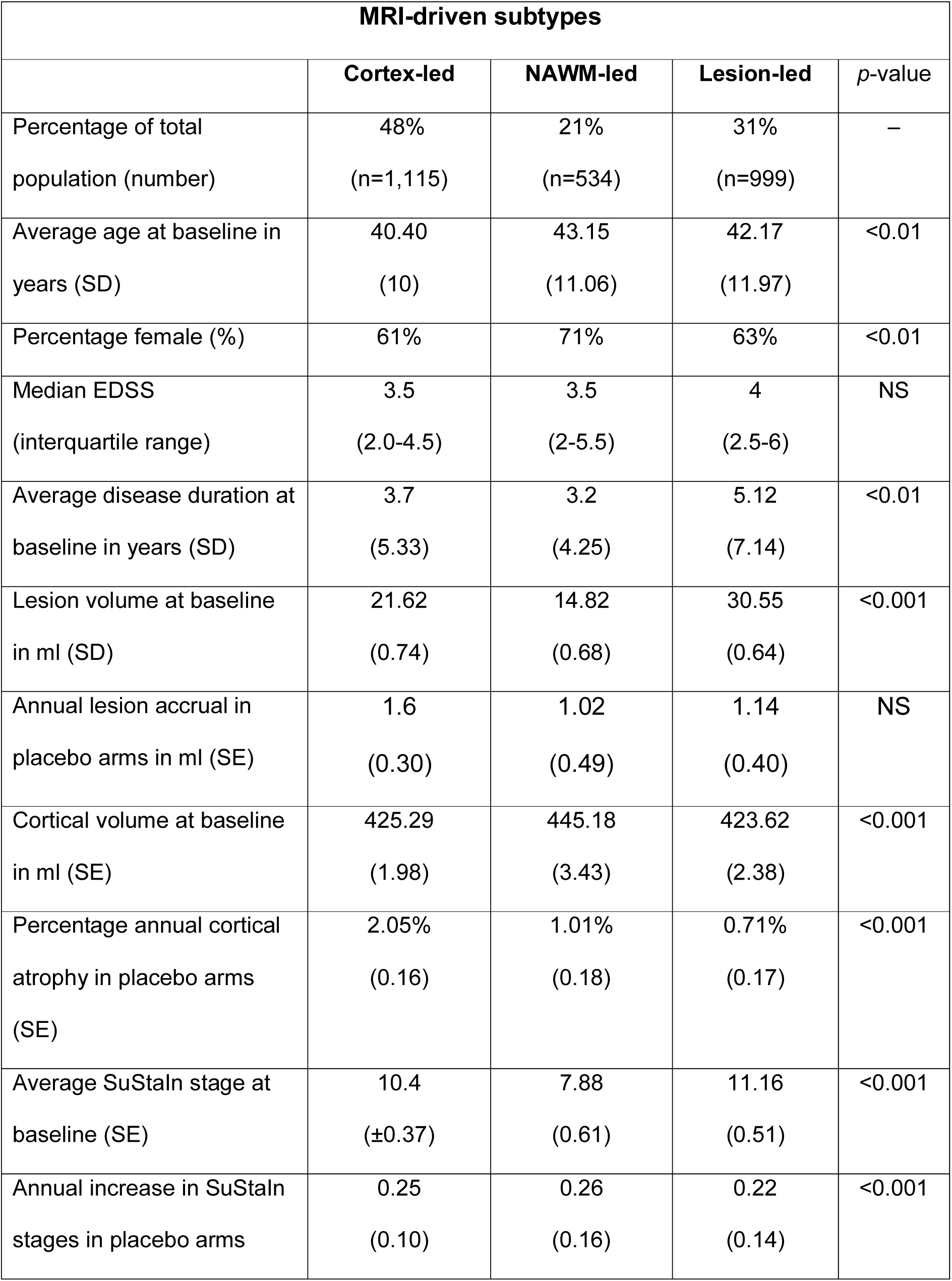
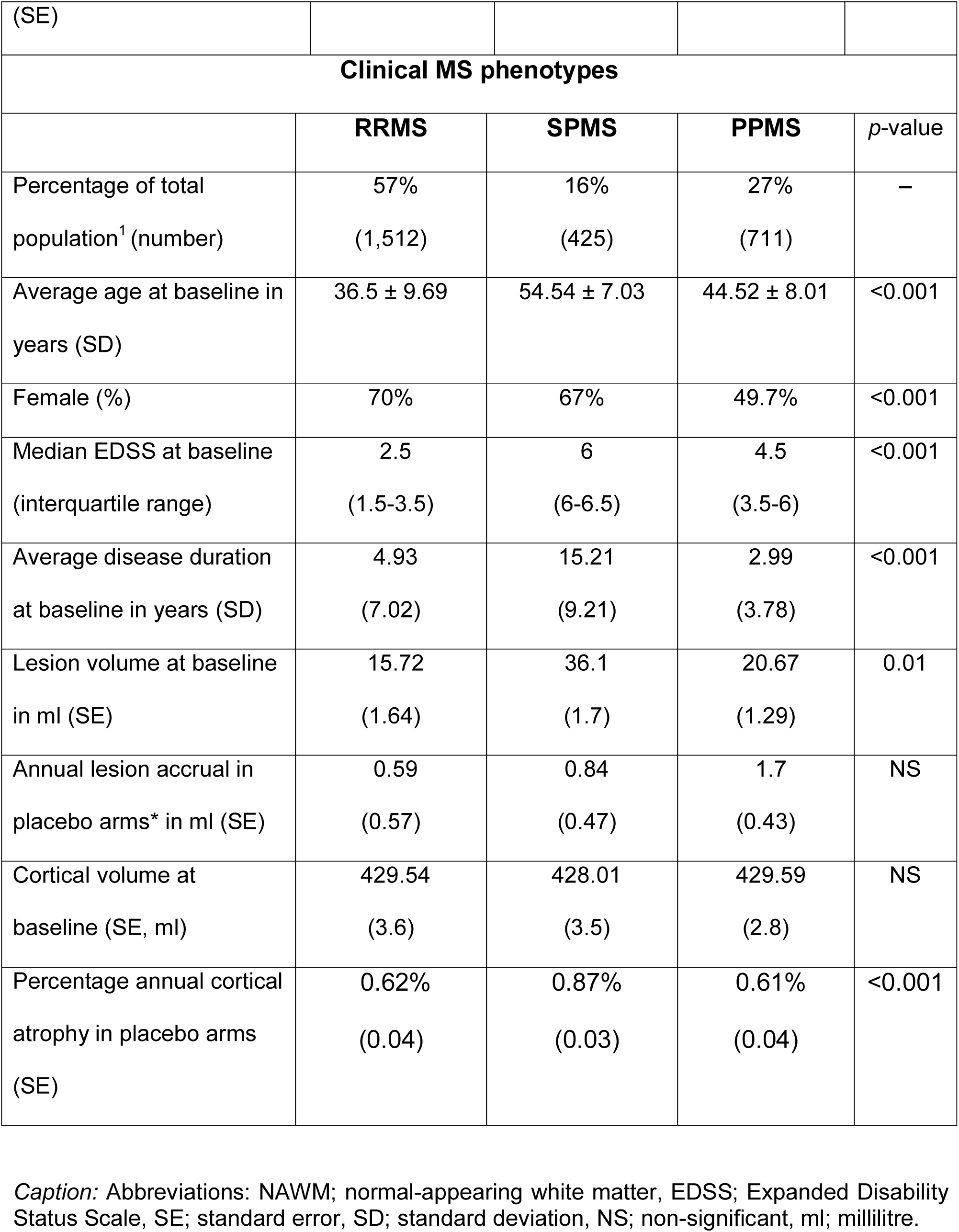
Demographic, clinical, and radiological characteristics of three MRI-driven subtypes and the clinical MS phenotypes in the external validation set.

When exploring the standard MS phenotypes in the external validation set (**Table 3**), as expected SP and PP MS patients were older and had higher EDSS than RRMS. SPMS had the highest lesion load at baseline, but a numerically lower cortical volume than RRMS and PPMS, though the difference in cortical volume was not statistically significant. The rate of cortical atrophy was significantly faster in SPMS than RRMS and PPMS (**Table 3**).

When exploring the distribution of MRI-driven subtypes across the MS phenotypes, the cortex-led was the most common subtype in RRMS (46.7%), and PPMS (50%) but the least common subtype in SPMS (11.1 %). The lesion-led was the most common subtype in SPMS (60.5%) and the second most common subtype in RRMS (33%) and PPMS (32.9%). The NAWM-led was the second most common subtype in SPMS (28.3%), but the least-common in PPMS (16.6%) and RRMS (19.8%).

### Differences in the risk of disability progression between the MRI-driven subtypes in the external validation set

When we investigated the differences in the risk of developing 24-week-confirmed disability progression (CDP) across the three subtypes–which were detected at *baseline*–within each dataset of the external validation set, we found that the lesion-led subtype was consistently associated with the highest risk of CDP, except for the MS-SMART RCT. In the BRAVO trial, there was a statistically significant difference in reaching 24-week-CDP among the three subtypes (log-rank test for three-group comparison, *p*=0.003) (**Figure 4a**). The lesion-led subtype had 82% (95% confidence interval: 48.6% to 115%, p<0.001) higher risk of reaching the 24-week CDP than the cortex-led subtype (**Figure 4a**). There were no differences between lesion-led and NAWM-led subtypes, and between cortex-led and NAWM-led subtypes.

**Figure 4.**
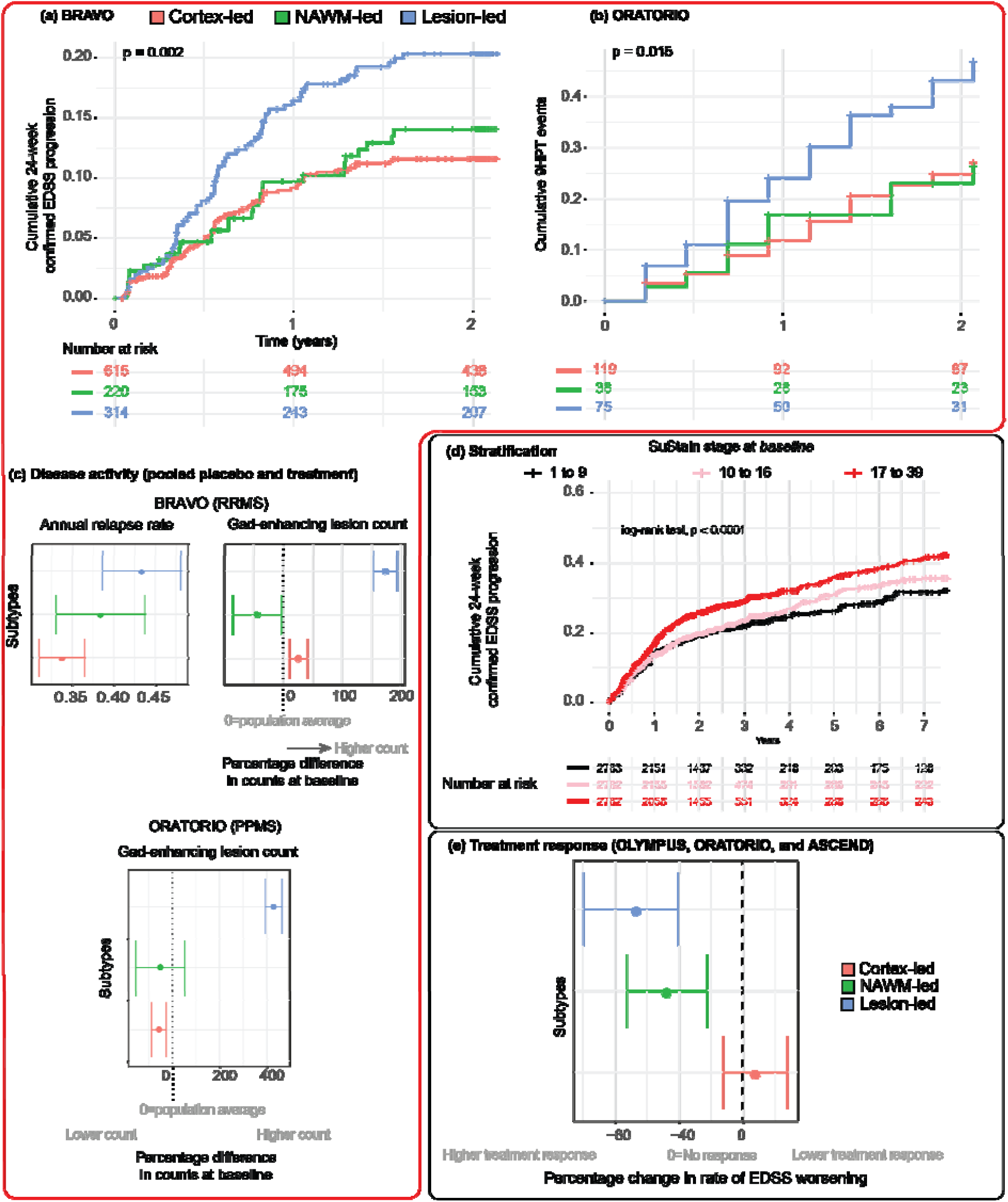
Predicting disability progression, disease activity, and treatment response. this figure shows our analyses in the external validation sets (red border) and pooled datasets of train and test sets (black border). (a) Patients in the lesion-led subtype had a faster EDSS progression in BRAVO; (b) patients with lesion-led subtype had a faster 9-Hole Peg Test progression (see the main text for definition) compared to the other two subtypes in the placebo arm of the ORATORIO trial (we did not use treatment arm to look at longitudinal changes here because it was a positive trial). The vertical axis shows the 24-week confirmed EDSS progression; (c) top diagram: the lesion-led subtype had more annual relapse rates when looking at all patients in the BRAVO trial than the NAWM-led or cortex-led subtypes; bottom diagram: the lesion-led subtype had more gadolinium enhancing lesions in all patients of ORATORIO trial at baseline. Error bars represent the standard error; (d) this section shows that patients who were in the higher tertile of SuStaIn stages had a shorter time to progression: the higher the stage at baseline, the shorter the time to reach 24-week confirmed EDSS progression. When we repeated this analysis inside each subtype we found similar results (these are not shown); (e) Shows the change in EDSS worsening in MRI-driven subtypes in the pooled treatment arms of the ORATORIO, ASCEND and OLYMPUS trials compared to the corresponding subtypes in the pooled placebo arms (e.g., lesion-led subtype on treatment *vs*. lesion led subtype on placebo and so forth). Patients in the lesion-led subtype had the largest reduction in the rate of EDSS worsening and were the only group who had a significant treatment response. Error bars represent the standard error. Acronyms: 9HPT, 9-Hole Peg test; NAMW, normal-appearing white matter; EDSS, Expanded Disability Status Scale; RRMS, relapsing-remitting multiple sclerosis; PPMS, primary progressive multiple sclerosis.

Similarly, to the BRAVO results, in the ORATORIO trial, there was a significant difference in reaching 24-week-CDP across the three subtypes (log-rank test for three-group comparison, *p*=0.015) on the placebo arm. The lesion-led subtype had 99% higher risk of developing a 20% deterioration on the 9-Hole Peg Test (95% confidence intervals=19% to 231%, p=0.007) than the cortex-led subtype (**Figure 4b**); there were no differences between lesion-led and NAWM-led subtypes, and between cortex-led and NAWM-led subtypes.

In the MS-SMART trial, the lesion-led subtype had on average 3.35 lower PASAT score at baseline than the cortex-led subtype, though this was not statistically significant. Over time the lesion-led subtype had the fastest average decline in the Paced Auditory Serial Addition Test or PASAT, which was significantly faster than the cortex-led subtype (difference in rate= −2.10, 95% CI= −0.26 to −3.93, *p*=0.03) but this rate was not significantly different from NAWM-led subtype. There were no statistically significant differences across three subtypes in the CDP, 9-hole peg and timed walk tests performance.

In the CLIMB observational cohort, patients with the lesion-led subtype had a significantly shorter time to reach 24-week CDP than the other two subtypes (log-rank test for three group comparison, p=0.035) **(Supplementary Figure 3**). The lesion-led subtype had 75% higher risk of reaching 24-week-CDP than the NAWM-led subtype (95% confidence intervals=14% to 170% higher risk, p=0.01). There were no significant differences between lesion-led and cortex-led subtypes and between NAWM-led and cortex-led subtypes.

### Differences in disease activity between the three MRI-driven subtypes in the external validation set

When we investigated the differences disease activity (i.e., relapse rate for BRAVO and MS-SMART and enhancing lesions for BRAVO and ORATORIO, chosen according to data availability) between subtypes within each dataset of the external validation set, we found that the lesion-led subtype consistently showed the highest disease activity. In MS-SMART there were no statistically significant differences in relapse rate across the subtypes. In the BRAVO trial, the lesion-led subtype had a higher annualised relapse rate (average=0.43, 95% confidence intervals=0.35 to 0.51) than the cortex-led subtype (average=0.33, 95% confidence intervals=0.29 to 0.37; *p*=0.04) (**Figure 4c)**.

When looking at gadolinium-enhancing lesion counts in the placebo and treatment arms of the BRAVO at baseline, the lesion-led subtype had more gadolinium-enhancing lesions (average=3.44 lesions, interquartile range=3) than the cortex-led subtype (average=1.2, interquartile range=1, Poisson model-estimated difference=146%, standard error=20.5%, *p*<0.001) (**Figure 4c)**. The NAWM-led (average=0.72, interquartile range=1) and the cortex-led subtypes did not show different numbers of gadolinium-enhancing lesions (p=0.1) (**Figure 4c)**. Similarly, when looking at gadolinium-enhancing lesions in both the placebo and treatment arms of the ORATORIO trial at baseline, the lesion-led subtype showed more lesions (average=2.25, interquartile range=2) than the cortex-led subtype (average=0.42, interquartile range=0, Poisson model-estimated difference between two groups=486%, standard error = 109%, p<0.001). Similarly, the lesion-led subtype had more gadolinium-enhancing lesions than the NAWM-led subtype (average lesion in the NAWM-led=0.21, interquartile range=0, Poisson model-estimated difference= 479%, p=0.001) (**Figure 4c)**. The NAWM-led subtype (average=0.42) and the cortex-led subtype (average=0.21) showed similar (p=0.3) numbers of gadolinium-enhancing lesions (**Figure 4c)**.

### Relationship between the MRI-driven subtypes and stages and both disability progression and treatment response

When looking at stages of the MS subtypes in all the available datasets together, patients who had the highest tertile of SuStaIn stage at baseline (from 17 to 39) had the shortest time to 24-week-CDP (independent of subtype, log-rank p<0.0001) (**Figure 4d**). Average (95% confidence interval) risk of developing 24-week-CDP at any particular time was 37% (22-53%) higher in this group with respect to the group with the lowest tertile of stage (from 1 to 9) (p<0.0001) and 30% (17 to 46%) higher (p<0.0001) in this group with respect to the middle tertile group (from stage 10 to 17) (**Figure 4d**).

Additionally, there were significant associations (standardised *β*) between MRI-driven subtypes (overall subtype effect, *β*= 0.04, standard error= 0.01, p=0.02) and stages at baseline (*β*= −0.06, standard error= 0.02, p<0.001) with the time-to-24-week-confirmed EDSS progression. There were no significant associations between the standard clinical phenotypes (overall effect across RRMS, SPMS and PPMS *β*= 0.18, standard error= 0.15, p=0.22) or baseline EDSS (*β*=0.02, standard error = 0.03, p=0.26) with the time-to-24-week-CDP.

### Consistency of the subtype membership over time

Presented results so far have been on subtypes detected using the baseline MRI scans. We looked at longitudinal stability of subtype membership, too, to examine the reliability of SuStaIn. When we looked at the first and last visit of 4,741 patients who were assigned to one of the three MRI-driven subtypes with probability of more than 99% at baseline, 95.9% stayed in the same subtype over time (277 patients changed subtype: 37 from cortex-led to NAWM-led, 59 from cortex-led to lesion-led, 64 from NAWM-led to cortex-led, 4 from NAWM-led to lesion-led, and 32 from lesion-led to cortex-led, and 1 from lesion-led to NAWM-led). When including all subjects without a threshold of probability, 75% remained in the same subtype.

### Difference in treatment response between the three MRI-driven subtypes

When we tested whether there were differences in *treatment responses*–defined as the difference for each subtype in EDSS worsening on treatment *vs* placebo–we found that that the lesion-led subtype was the only subtype showing a significant treatment response in three phase 3 RCTs in SPMS and PPMS trials together (ORATORIO, ASCEND, and OLYMPUS^5,13,14^) (**Figure 4e)**. Patients with the lesion-led subtype on treatment showed a significantly slower worsening of EDSS than the same subtype on placebo (percentage average difference ± standard error: −66% ± 25.6%, p=0.009). The NAWM-led and cortex-led subtypes did not show a slower worsening on treatment than the same subtype on placebo (−22% ± 25%, p=0.06; 27.6%, ± 20.17%, p=0.7, respectively) (**Figure 4e**).

Similarly, in the pooled analysis of RRMS (OPERA1, 2, and DEFINE/CONFIRM/ENDORSE) the lesion-led subtype on treatment showed a significant reduction in the rate of EDSS worsening compared to the same subtype on placebo, or active comparator arms (−89% ± 44%, p=0.04). There was no significant reduction for the NAWM-led (−75% ±198%, p=0.74) or the cortex-led subtype (63%, ±164%, p=0.70) when compared to the same subtype on comparator arms.

## Discussion

The MRI-driven subtyping of MS, based on a recent innovation in unsupervised machine learning (SuStaIn), demonstrated distinct patterns of accumulation of abnormality on MRI over time in the train and internal validation set and in the external, independent validation set. We identified three MRI-driven subtypes: 1) Cortex-led, 2) Normal-appearing white matter (NAWM)-led, and 3) Lesion-led. The cortex-led subtype was the most common subtype in both the internal and external validation sets; it showed early cortical atrophy, with slower accrual of lesions over time than the lesion-led subtype in the external validation set. The lesion-led subtype was the most common subtype in the SPMS phenotype in the train and internal validation set (no SPMS patients were in the test set) and showed a longer disease duration than the other two subtypes; additionally, it showed an early accumulation of lesions. The NAWM-led showed early reduction in T1/T2 ratio with slower accrual of lesions than the lesion-led subtype, and slower cortical atrophy than the other two subtypes. These MRI-driven subtypes showed high consistency (Bhattacharyya coefficient range=0.94 to 0.96) across 14 datasets (and MRI protocols) and, over time, patients who were assigned to a subtype with high (99%) certainty, rarely (4.1%) transitioned into a different subtype. We tested our model in external datasets and with variables that we did not use in training, such as relapses, gadolinium-enhancing lesions, EDSS progression, 9 Hole Peg Test performance, and PASAT; We found that MRI-driven subtypes predicted disability progression, treatment response, disease activity (relapse rate and gadolinium-enhancing lesions) and cognitive decline. When we stratified patients according to the SuStaIn stage, we found that higher stages at baseline could predict faster disability progression.

The lesion-led subtype was the least common (25% of patients) in the train and internal validation set and the second most common (31% of patients) in the external validation set. In both sets, the most common subtype overall was the cortex-led. In the RRMS phenotype of both sets, a cortex-led subtype was more common than a lesion-led subtype. In SPMS of both internal and external sets, more patients were classified as having a lesion-led subtype than a cortex-led one. A faster EDSS progression in the lesion-led subtype explains why there were more lesion-led patients with the SPMS phenotype. Given that not all those with RRMS will develop SPMS, RRMS patients are a combination of those less likely (cortex-led) and more likely (lesion-led) to eventually develop SPMS. SuStaIn is a probabilistic model; so it enabled us to group patients across a spectrum of three subtypes (Figure 3). Although when we applied SuStaIn to follow-up scans the majority of patients remained in the same subtype, further studies with longer follow-ups are required to clarify whether these subtypes are distinct entities. Our modelling nonetheless provides a powerful tool to stratify patients in clinical trials using baseline scans.

Our findings provide insights into the sequence and focus of pathology, albeit with the caveat that MRI measures are not pathologically specific. The cortex-led had early cortical atrophy and lower lesion volumes than the lesion-led subtype. Cortical atrophy in the cortex-led subtype started posteriorly and spread forward, increasingly involving deep grey matter, with abnormalities in NAWM occurring late and with slower lesion accumulation. Our findings suggest a pathological process that is predominantly in the cortex rather than WM-based, dominated by posterior and then more extensive neurodegeneration (as reflected by atrophy)^15,16^. In contrast, the lesion-led subtype starts with marked accumulation of lesions, severe early atrophy in the deep grey matter structures, and then cortical atrophy, with NAWM abnormalities as a late feature. The ongoing cortical atrophy in the placebo arms was faster in the lesion-led subtype than the cortex-led or NAWM-led subtypes. This finding suggests a process more closely linked with WM lesion accrual. These findings are consistent with neurodegeneration in deep grey matter secondary to lesion accumulation^15,16^. Similarly, faster cortical atrophy in this subtype may be secondary to white matter lesion effects on tracts, or concomitant inflammatory processes in white matter and grey matter^17,18^. The sequence of the NAWM-led subtype suggests a more diffuse process, with limited WM lesion formation even by the late stages, and subsequent GM abnormalities detectable anteriorly and then posteriorly.

### Associations between MRI-driven subtypes and clinical outcomes

MRI-driven subtyping has the potential to predict treatment response and disability progression irrespective of clinical phenotypes. Patients with the lesion-led subtype had worse disability outcomes and a more active disease (clinical and radiological) than the other two subtypes. The MRI-driven subtypes and stages were more strongly associated with EDSS worsening than the baseline EDSS or clinical phenotypes. We confirmed these results in the external validation set, where the baseline EDSS ranged from 3.5 to 4.0 for all the three MRI-driven subtypes; however, the MRI-driven subtypes still predicted disability progression and cognitive decline (EDSS, 9HPT performance, and PASAT across different data sets). These findings suggest that our newly identified subtypes may have a better prognostic value than the clinical phenotypes of MS. The lesion-led subtype was the only subtype that showed a significant treatment response in both RR and progressive (PP and SP) MS trials. Our work confirms previous observations that current clinical MS phenotypes do not have clear underpinning biological basis^2,3,6^ but also provides new subtypes which are likely to share pathogenic mechanisms. Our results in the lesion-led subtype suggests that the inflammatory pathological processes are likely to be the dominant driver of cortical atrophy, lesion accumulation, and disability worsening because the lesion-led subtype had a greater reduction in EDSS worsening (treatment response) than the other two subtypes after treatment exposure. Since our MRI-driven subtyping can work also using MRI from a single time-point, it has the potential to prospectively enrich trials with those most likely to respond to treatment.

### Limitations and future directions

Changes in normal appearing white matter relate to diffuse pathological processes in MS and have important clinical implications^19^. Yet it is not feasible to measure such changes with more advanced modalities such as the diffusion tensor and magnetisation transfer indexes in phase 3 clinical trials which involve tens of countries and sites. T1/T2 ratio measures in the normal appearing white matter is considered a “crude” measure but is the only (semi) quantitative measure that is both universally available and is informed by the microstructural changes. Since this measure can be affected by the choice of MRI protocol and scanner, we paid special attention to “trial” and “centre” effects separately. To mitigate potential differences across scanners we used an internal reference (ventricles) to normalise values (see Supplementary Methods). There were 728 centres in this study; when we looked at the “centre” or “site” effects and compared it with the “subtype” effect across MRI variables (including T1/T2), the MRI measures were more strongly related to subtype than centre. To measure and examine trial effects, we used leave-one-dataset out cross-validation and found excellent consistency across cross-validation folds. Our model could predict clinical and cognitive outcomes (EDSS progression, 9-hole peg test, PASAT) when applied to new centres in unseen data sets, which confirms that the centre effects are unlikely to significantly affect the predictive performance.

We have identified MRI-driven subtypes using clinical trials whose image quality is high. Further work is required to demonstrate MRI-driven subtyping in real-world clinical data. Additionally, the analysis pipeline that identifies the subtype requires imaging expertise and is not yet feasible for use outside research centres, although its implementation in treatment trials is achievable because MRI in being used in screening of recent clinical trials^20^. Spinal cord is affected from early stages in MS and its atrophy is associated with disability^21^. However, spinal cord data is not routinely acquired in MS trials and was not available in our study. Future studies with spinal cord data should investigate whether spinal cord measures can contribute to SuStaIn subtyping and staging, and whether they do so independently or concurrently with brain atrophy. Another aspect for future studies will be to investigate pathological underpinnings of these MRI-driven subtypes.

## Conclusion

We used SuStaIn to obtain MS subtypes and stages based on distinct patterns of accumulation of MRI abnormalities, thereby reflecting the pathogenic mechanisms of MS better than phenotypes based on purely clinical descriptions. MRI-driven staging can predict disability and MRI-driven subtyping has the potential to be used prospectively to enrich clinical trials with patients who are more likely to respond to treatments.

## Methods

### Participants

We collected clinical and MRI data from 15 MS randomised-controlled trials (RCTs): five trials of PPMS^5,13,22,23^, six trials of SPMS^14,24–28^, and four trials of RRMS^29–31^; we also included three observational cohorts with mixed MS subtypes^5,13,14,22–27,29–36^ (**Table 1**).

Each RCT and observational study had received ethical approval and participants had given written, informed consents at the time of data-acquisition. The Institutional Review Board at the Montreal Neurological Institute (MNI), Quebec, Canada approved this study (Reference number: IRB00010120). The pharmaceutical companies who provided the fully anonymised, individual patient raw data, agreed to pooling data but not re-testing treatment response in individual RCTs.

We also included two datasets of healthy controls by downloading unprocessed MRI data from: (1) The S1200 Open Access release of the Human Connectome Project, and (2) The UK Biobank data, which were available for download on 1^st^ of February 2019. This project was approved by the UK Biobank (Reference number: 47233).

### Clinical outcomes

The Expanded Disability Status Scale (EDSS)^37^, which rates neurological impairment, was scored as per individual study protocol. The EDSS was obtained at least one month after a protocol-defined relapse. We defined disability progression confirmed at 24 weeks (or confirmed disability progression (CDP)) as a worsening of EDSS that was sustained on subsequent visits for at least 24 weeks. EDSS progression was defined as a ≥1.5-point increase from a baseline EDSS of 0, a ≥1-point increase from a baseline score of 0.5 to 5.5, and a ≥0.5-point increase from a baseline score greater than 5.5.

### Brain MRI Protocol

We collected the following brain MRI sequences: T1-weighted, T2-weighted, and Fluid Attenuated Inversion Recovery (FLAIR) MRI in all except three datasets (see **Supplemental Material** for details). We used brain 2D or 3D T1-weighted scans to segment grey and white matter tissues, FLAIR and T1-weighted scans to segment lesions, and T2-weighted scans, together with T1-weighted scans, to obtain T1/T2 ratio. Details of MRI protocols are explained in publications associated with each dataset^5,13,14,22–27,29–33,35,36,38^.

### Image processing

We processed MRI scans from MS and healthy volunteers to obtain the following 18 variables according to the Neuromorphometrics atlas (http://www.neuromorphometrics.com):

- Volumes of the bilateral frontal, parietal, temporal, and occipital grey matter, limbic cortex, cerebellar grey matter and white matter, brainstem, deep grey matter and cerebral white matter
- Volume of total T2 lesions
- Regional T1/T2 ratio of normal-appearing white matter in the corpus callosum, frontal, temporal, parietal, and occipital lobes, cingulate bundle and cerebellum.

Details of image analysis and quality control pipelines are explained in detail in the **Supplementary Material**.

## Statistical analysis

### Outline

The analysis was divided in two parts. In the first part, we carried out the same data processing previously described^9^, using SuStaIn. We first trained and internally validated (cross-validated) SuStaIn in a “train and internal validation” set, and in the second part we tested it by using an “external” (unseen and independent) set. We also investigated the associations between SuStaIn subtypes and stages (and standard clinical MS phenotypes) and both disability progression and treatment response by using all the available datasets. In the second part of the analysis, we explored whether there were differences in treatment responses between MRI-driven subtypes in three phase 3 RCTs in RRMS and in three phase 3 RCTs in progressive MS.

#### (1) Model training, internal and external validation

The first part of the analysis included the following steps (**Figure 1**):

1. Adjusting MRI measures for nuisance variables.
2. Calculating Z-scores of MRI measures based on healthy controls.
3. Separating, *a priori*, MS trials and cohorts into a “train and internal validation” set for model training and validation, and an “external” test set for model testing.
4. Selecting MRI variables in the “train and internal validation” set.
5. Identifying the optimal model by carrying out the leave-one-dataset-out cross-validation
6. Testing the newly developed model on the external validation set, thereby confirming its Generalisation ability and investigating the differences in the risk of progression and disease activity between subtypes.
7. Investigation of the relationship between subtypes and stages and both disability progression and treatment response using all the available datasets together (train and external sets).
8. Investigation of the reliability and stability of the model using all the available datasets together.

We now explain each step below:

##### 1) Adjusting MRI measures for nuisance variables

For each of the 18 MRI variables listed above, we used the two datasets of healthy controls together to fit a Bayesian linear regression model with the total intracranial volume, sex, age and age squared as independent variables, and each MRI variable as the outcome. We calculated the expected values for each visit using this model and subtracted the observed values to obtain residual values of each MRI variable. We refer to the residual values as “adjusted values”. We used BAS package version 1.5.3 and R version 3.6.0^39^. We evaluated “study” and “centre” effects separately, as explained below in the internal validation and in the Supplemental Methods.

##### 2) Calculating Z-scores of adjusted MRI measures based on healthy controls

We calculated the Z-scores for each MRI variable at each participant’s visit by subtracting the adjusted mean value of the healthy volunteers from the adjusted observed value in patients and dividing each patient’s MR variable by the standard deviation of the healthy volunteers.

##### 3) Separating, a priori, MS trials and cohorts into a “train and interval validation” set and an “external” test set

From the 18 datasets available, we *a priori* chose 14 datasets to create a “train and internal validation” set, which was used for model training and internal validation (or cross-validation). These were three phase 3 RRMS trials^29,30^, three phase 3 PPMS trials^5,22,23^, two phase 3 SPMS trial^14,27^, three phase 2 SPMS trials^24–26^, one phase 2 PPMS trial^40^, and two observational cohorts^35,38^ (see **Table 1** and **Figure 1** for the complete list).

We set aside the remaining four datasets to create an external validation set, which was used to perform the model testing: one phase 3 RCT in RRMS^31^, one phase 3 RCT in PPMS^13^, one phase 2 RCT in SPMS^28^, and one observational cohort with mixed MS subtypes^36^ (**Table 1**).

##### 4) Selecting MRI variables in the “train and internal validation” set

To reduce the dimensionality of the models and computational expenses, we selected the MRI variables which were found to be ‘abnormal’ in MS when compared with healthy controls. To do so, we carried out pairwise comparisons between healthy volunteers and patients at their baseline visit and selected the MRI measures whose differences between the groups were associated with a moderate to large effect sizes (>0.5 Cohen’s D effect size).

##### 5) Identifying the optimal model by carrying out the leave-one-dataset-out cross-validation

We entered the MRI variables resulting from the previous step into SuStaIn. Since lower values of volume and T1/T2 ratio are expected to be associated with increased disability, we flipped their signs so that higher Z-scores and estimated stages represented disease worsening.

To find the optimal model (that is the model with the highest likelihood of explaining MRI variables), we carried out the internal validation with leave-one-dataset-out cross-validation, which allowed us to choose the number of MS subtypes, quantify the uncertainty associated with a given subtype “trajectory” (or the evolution of MRI abnormalities), and evaluate the stability and robustness of the model across different trials, and MRI protocols. With the leave-one-dataset-out cross-validation procedure, we trained the model on 13 out of the 14 datasets, and evaluated it using the remaining dataset (*held-out* sample). We permuted the training and held-out samples until every dataset was used once as held-out sample, thereby iterating the procedure 14 times. The steps were as follows (**Figure 1**):

I. We selected the best fitting model, which means the model with the optimal number of subtypes, and sequence of MRI abnormality changes in the same subtypes. We started by fitting the SuStaIn model on the 13 training folds with only one subtype and then increased the number of subtypes in steps of one. We calculated the log-likelihood (which expresses predictive accuracy) of each held-out fold for each model and chose the fitted model with the number of subtypes that maximised this log-likelihood.
II. We estimated the uncertainty of the quantification (posterior distribution) using the Markov Chain Monte Carlo (MCMC) algorithm with 100,000 iterations to sample from the posterior distribution of the most likely sequences found in the previous step.
III. To evaluate how consistent MRI-driven subtypes were across these 14 datasets in the “train and internal validation set”, we quantified the effects of dataset (and therefore MRI protocol) on subtype trajectory. To do so, we quantified the degree of overlap of posterior distributions of sequences for each subtype across 14 iterations of cross-validation. For this purpose, we used the Bhattacharyya coefficient^41^ between each pair of subtypes from different folds. The Bhattacharyya coefficient ranges from 0 (no agreement) to 1 (perfect agreement). We calculated all pairwise Bhattacharyya distances across all folds and subtypes pairs for the optimal model and reported the average and standard deviation for each subtype.
IV. We fitted our final trained model on all the 14 datasets in “train and internal validation” set to obtain the optimal model to be used for external validation. We obtained MCMC samples and visualised the uncertainty of the final model with its posterior distribution.

To compare gender frequencies among MRI-driven subtypes and clinical phenotypes, we used Chi-square test. To compare ordinal and continuous outcome variables (e.g., EDSS, SuStaIn stages, age and disease duration), and lesion and cortical volumes we used general linear models (with Poisson distribution for ordinal variables). For longitudinal analysis of lesion volume and cortical volume we used mixed-effects models. In these mixed-effects models, we included hierarchical random effects: the visit variable was nested in ‘subject’, and ‘subject’ variable nested in the ‘dataset’ variable; we only used the placebo arms of the RCTs to evaluate the natural course of MRI-driven subtypes in the absence of treatments and included total intracranial volume, age and sex as fixed-effects, nuisance variables.

##### 6) Testing the newly developed model on the external validation set, thereby confirming its Generalisation ability, and investigating the differences in the risk of progression and disease activity between subtypes

We tested our trained model on the external validation set, which was not used for model training and internal validation. As mentioned above, this external validation set included one phase 3 RCT in RRMS (BRAVO)^31^, one phase 3 RCT in PPMS (ORATORIO)^13^, and one observational study with mixed MS subtypes (CLIMB)^36^, and one phase 2 RCT in SPMS (MS-SMART^28^) (**Table 1**).

First, we applied SuStaIn to the whole test set to obtain subtype membership for each subject’s visit at *baseline* (study entry). Secondly, to investigate the differences in the hazard ratio of reaching the 24-week-CDP between the MS subtypes within each trial, Cox regression models were used. In the BRAVO RCT and CLIMB study, we performed a survival analysis to calculate the time-to-24-week CDP of each MRI-driven MS subtype. In the phase 3 RCT in PPMS (ORATORIO^13^), we considered only the placebo arm for the longitudinal analysis because the time-to-event analysis was modified by the experimental drug^13^. Since the number of 24-week confirmed EDSS progression events per MS subtype in the placebo group of the ORATORIO trial was too small (<23 events per subtype on average) to provide statistically reliable results, we defined an event as ≥20% increase in the averaged 9 Hole-Peg Test time between two hands, as suggested by a previous study^42^. Thirdly, to investigate whether there were differences in disease activity between the MRI-driven subtypes, we used the annual relapse rate in the BRAVO RCT (in the ORATORIO trial relapses were extremely rare) and the number of gadolinium-enhancing lesions at baseline in the BRAVO and ORATORIO trials, previously reported^13,31^. We used a Poisson model (with Poisson family distribution in R) in which the lesion count at baseline in all patients (placebo and treatment arms merged together) was the outcome and the MRI-driven subtype was the independent variable. The CLIMB study did not include information on relapse rate and gadolinium enhancing lesion count. MRI data with gadolinium injection were not acquired in MS-SMART.

##### 7) Investigating relationship between SuStaIn subtypes and stages and both disability progression and treatment response using all the available datasets together (train and test sets)

We used SuStaIn to estimate subtype stages along a trajectory or a ‘sequence’. Since there were 13 variables with three Z-scores each, each subtype included 39 stages, which ranged from one (the earliest stage) to 39 (the last stage). To investigate whether the MRI-driven subtypes and the standard clinical MS phenotypes were associated with disability progression, we constructed a mixed-effects model. In this model, time to reach 24-week-confirmed EDSS progression was the outcome variable and ‘trial’ was a random-effects variable. Fixed-effects predictors were MRI-driven subtypes and stages at baseline, standard MS phenotypes, age, sex, and EDSS at baseline.

##### 8) Investigation of the reliability and stability of the model using all the available datasets (train and test sets)

We performed additional analyses to test the reliability and stability of the SuStaIn subtypes over time in both the train and test set (see **Supplemental Material** for details).

#### (2) Difference in treatment response across MRI-driven subtypes

In the second part of the analysis, we explored whether there were difference in treatment responses between the MRI-driven subtypes, by looking at the rate (or slope) of EDSS worsening in three phase 3 RCTs in progressive MS (ORATORIO, ASCEND, and OLYMPUS^5,14,42^) pooled together, and in three phase 3 RCTs in RRMS (DEFINE-CONFIRM-ENDORSE, OPERA1, and OPERA2) also merged together. We chose these trials because they were either positive trials^13,30,43^ or had a subgroup that showed a trend towards a treatment response in previous publications^5,14^. For the RRMS trials, we merged the arms with different doses of the experimental drug, included the placebo arms, and excluded the active comparator arms. We used a linear mixed-effects model in which EDSS was the outcome variable with ‘group’, time, and group × time interaction as the independent variables. ‘Group’ was a binary variable indicating either a given subtype on treatment or the same subtype on placebo. To adjust for repeated measures and correlated residual errors, we added hierarchical random effects to our model, in which visits were nested in the ‘subject’ variable. We reported the difference in percentage change of EDSS worsening between groups, which we refer to as ‘treatment response’ throughout this manuscript.

We used NLME package version 3.1 and Survival package version 2.44 inside R version 3.6.0 for statistical analysis^44,45^.

## Data Availability

Data in the form of CSV files are available from the main authors to reproduce the results of this study after publication.

## Acknowledgments

This study was supported by the International Progressive MS Alliance (IPMSA, award reference number PA-1603-08175). We are grateful to all the IPMSA investigators who have contributed trial data to this study as part of EPITOME: Enhancing Power of Intervention Trials Through Optimized MRI Endpoints network (see the list of investigators in the appendix). This study was supported by the National Institute for Health Research University College London Hospitals Biomedical Research Centre. O.C. is an NIHR Research Professor. We are grateful to Professor Geraint Rees for his comments. We thank Rozie Arnaoutellis, Istvan Morocz, and Caramanos Zografos for coordinating and organising this study. We thank Jonathan Steel for IT support during this work. This research in part has been conducted using the UK Biobank Resource under Application Number 47233. Data have also been provided in part by the Human Connectome Project, WU-Minn Consortium (Principal Investigators: David Van Essen and Kamil Ugurbil; 1U54MH091657) funded by the 16 NIH Institutes and Centres that support the NIH Blueprint for Neuroscience Research; and by the McDonnell Centre for Systems Neuroscience at Washington University. D.C.A. has received funding for this work from Engineering and Physical Sciences Research Council Grants M020533, M006093, and J020990, as well as the European Union’s Horizon 2020 Research and Innovation Programme under Grant Agreements 634541 and 666992.

## Supplemental material

### Brain MRI Protocol

MRI protocols differed between trials but inside each trial a unique MRI protocol has been used (except for the CLIMB study). We included brain 2D or 3D T1-weighted, fluid attenuated inversion recovery (FLAIR), and T2-weighted MRI scans. **Supplementary Table 1** shows the list of included trials with corresponding publications that reported details of MRI protocol.

In the CLIMB study, in which the MRI protocol had changed over time from 2D T1-weighted MRI to 3D, we only included more recent 3D T1 weighted MRI data.

## Supplemental Methods

### Image analysis

#### Brain MRI data handling

We checked and labelled the sequence of MRI scans by visually inspecting nine slices of each MRI scan (three axial, three sagittal, and three coronal slices) with equal slice intervals from the coordinates of the “centre of gravity” of each scan. We organised and uploaded MRI data to an XNAT server (version 1.7.4)^47^. We implemented our image analysis pipeline inside XNAT with Nipype version 1.1.4 to enable large-scale high-throughput computing^48^.

#### Regional brain volume calculation

We aimed to analyse scans to extract volumes of the grey matter regions according to an established brain atlas developed by Klein and Tourville (Neuromorphometrics, http://www.neuromorphometrics.com, see above for the list)^49^. We applied an identical cross-sectional pipeline (treating each visit independently) to all the visits of patients and healthy controls in which T1-weighted, FLAIR and T2-weighted MRI were available. We chose a cross-sectional, rather than a longitudinal image processing pipeline, to ensure that our subtyping models can be used prospectively in the real-world datasets in which (future) follow up data are not yet acquired. We adapted our established MRI analysis pipeline, which we had previously validated in clinical trials and observational cohorts as explained elsewhere in detail^50,51^. Briefly, it included intensity inhomogeneity correction of the T1-weighted MRI with ITK version 5.0 N4-bias field correction algorithm^52^, automatic segmentation of hyperintense lesions of the FLAIR sequence using the consensus (intersection) mask of two different methods (the regression based method in Lesion Segmentation Toolbox version 2.0.15^53^ and a deep convolutional neural network based method in DeepMedic version 0.7.1^54^, trained and validated previously with manual lesion masks from MS patients), rigid registration of FLAIR to T1-weighted MRI with co-registration of the FLAIR lesion masks to T1-weighted MRI using ANTs version 2.1.0, and lesion filling with NiftySeg version 1.0^55^. We segmented and parcellated the brain into Neuromorphometrics atlas regions on lesion-filled T1-weighted scans using the Geodesic Information Flows (GIF) software version 3.0^56^. We used a modified version of this pipeline for the Siena cohort, ARPEGGIO and lamotrigine trials which did not have FLAIR but whose investigators had provided manually delineated lesion masks.

#### T1/T2 ratio calculation of the normal-appearing white matter regions

Lesion masks or brain volumes do not provide any quantitative information on microstructural changes in the white matter. We therefore chose T1/T2 ratio as a measure of extra-lesional white matter changes, because T1 and T2-weighted MRI are widely available in clinical trials and clinical practice (as opposed to more advanced MRI sequences such as diffusion imaging or magnetisation transfer ratio). T1/T2 ratio is an extensively used measures of microstructural changes^57,58^. We adapted available pipelines from the Human Connectome Project to calculate T1/T2 ratio maps for all trials^59^. We corrected for intensity inhomogeneity in T1 and T2-weighted MRI scans with N4 bias field correction algorithm. Next, we rigid-registered T1 and T2-weighted scans in a symmetric space, such that both modalities equally underwent only one interpolation to minimise interpolation artefacts. We calculated the T1/T2 ratio and normalised its value against the average T1/T2 ratio in the ventricles with the co-registered ventricular masks obtained from the GIF segmentation (explained above). We extracted T1/T2 ratio from bilateral normal-appearing white matter regions (see above for list of regions) after we removed co-registered lesions segmented in FLAIR from the white matter regions, which we refer to as normal appearing T1/T2 ratio throughout this manuscript. Since the T1/T2 ratio in the grey matter regions were highly correlated with grey matter volumetric results, we did not include any T1/T2 ratio in the grey matter in our models.

#### Quality control

We developed a pipeline to check the quality of results of our pipeline by automatically generating 18 images from segmentation results, lesion segmentations, and registration results which we manually reviewed. We re-ran image analysis pipeline where we identified mis-registrations or faulty segmentations. We did not exclude any visit in clinical trials to perform an intention-to-treat analysis in individuals who met the minimal MRI criteria (which was availability of T1-, T2-weighted, and FLAIR).

### Supplemental Statistical Analysis Centre effects

From the 18 data sets in the train and external validation sets, 14 were multi-centre, which means that their MRI data were acquired by two or more scanners. To compare “centre” effects with “subtype” effects, using all data from train and external validation (test), we fitted hierarchical mixed effects models in which MRI variables were outcome, “centre” and “subtype” were predictors, and “study” was the random effects variable.

### Reliability and stability of SuStaIn models: longitudinal subtyping

In addition to subtyping patients at baseline, we trained our model on the baseline subjects and predicted the probability of subtype membership for the available patient visits over time (32,602 visits). We reported the number of subjects who preserved the subtype membership. To calculate the annual rate of change in SuStaIn stages for each data-driven subtype, we fitted a mixed-effects model in which the SuStaIn stage was an outcome variable and time was the independent variable (fixed effects). In these models to adjust for hierarchical repeated measures, we defined nested random effects in which ‘time’ variable was nested in the ‘subject’ variable. To calculate longitudinal cortical atrophy in each subtype we used a similar mixed effects model and log-transformed the cortical volumes to obtain the annual percentage volume change.

## Supplemental Results

### Defining the optimal number of subtypes: model selection

We fitted models that had one to five subtypes for 14 cross-validation folds (total of 70 models, **Figure 1**). We used Cross Validation Information Criteria (CVIC) to choose the most optimal number. The CVIC was the most optimal (minimum value across models up to five subtypes) for the three subtype-model (**Figure 2b**).

### Centre vs subtype effects: Subtype was more strongly associated with clinical and imaging outcomes than the centre

MRI and clinical data in train and external validation sets were acquired at 728 different centres. EDSS was more strongly associated with subtype than centre (difference in standardised *β*= 0.04, standard error = 0.009, p<0.001). Similarly, when looking at the 13 MRI measures, their standardised *β* coefficients were significantly larger than centre coefficients, which means that they were more strongly associated with subtype than centre (all p values < 0.001).

**Supplementary Figure 1.**
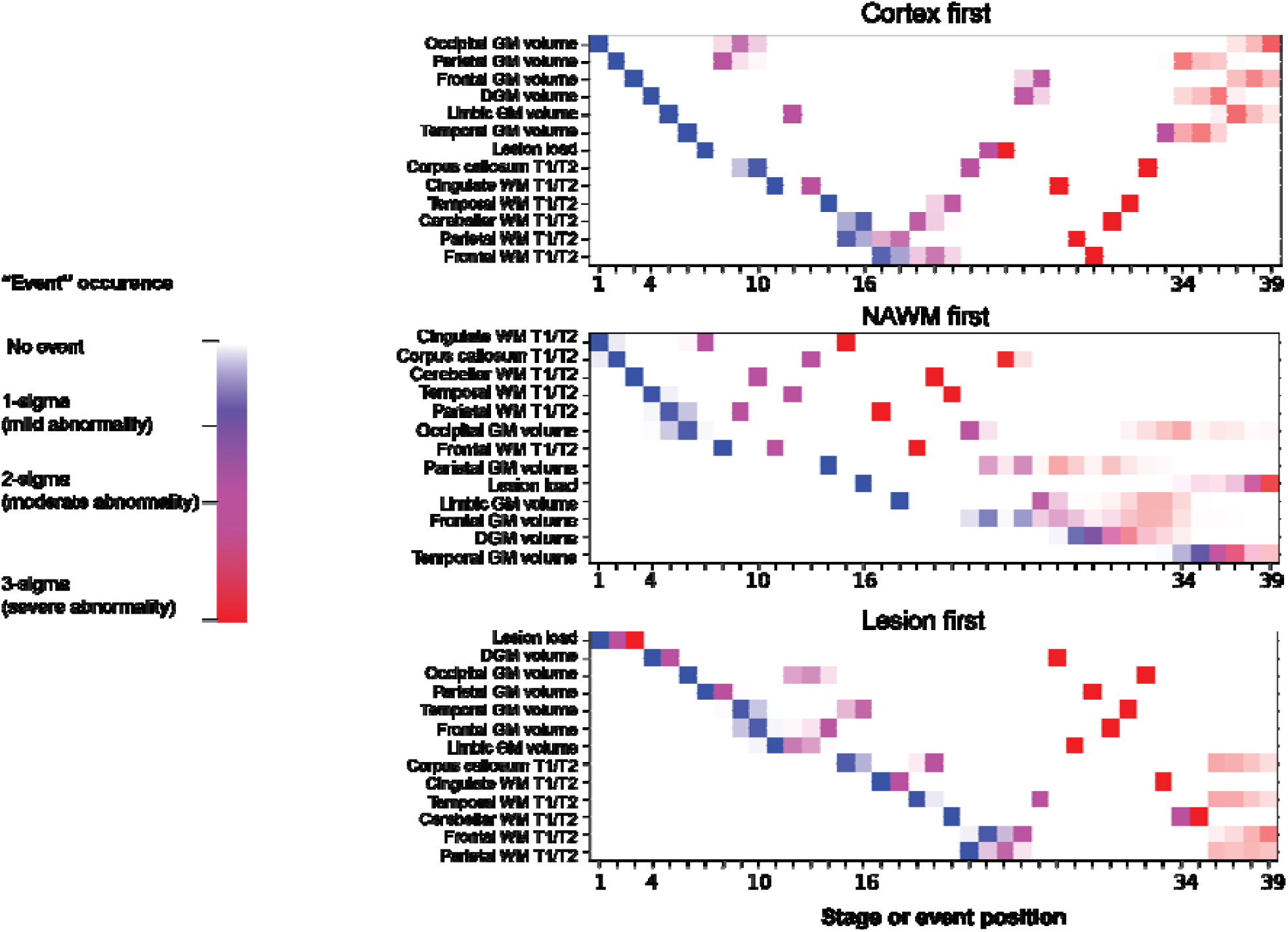
Positional variance diagram of three data-driven subtypes of multiple sclerosis. positional variance diagram for the three MRI-driven imaging subtypes. The three different colours represent the degree of abnormality based on Z-score (sigma or standard deviation) models: mild=blue, moderate=violet, and severe=red. The colour shades represent the uncertainty associated with each event position in the posterior distribution of 100,000 Markov Chain Monte Carlo samples. Acronyms: DGM, deep grey matter; T1/T2, T1-T2 ratio; WM, white matter; GM, grey matter; DGM, deep grey matter; NAWM, normal-appearing white matter.

**Supplementary Figure 2.**
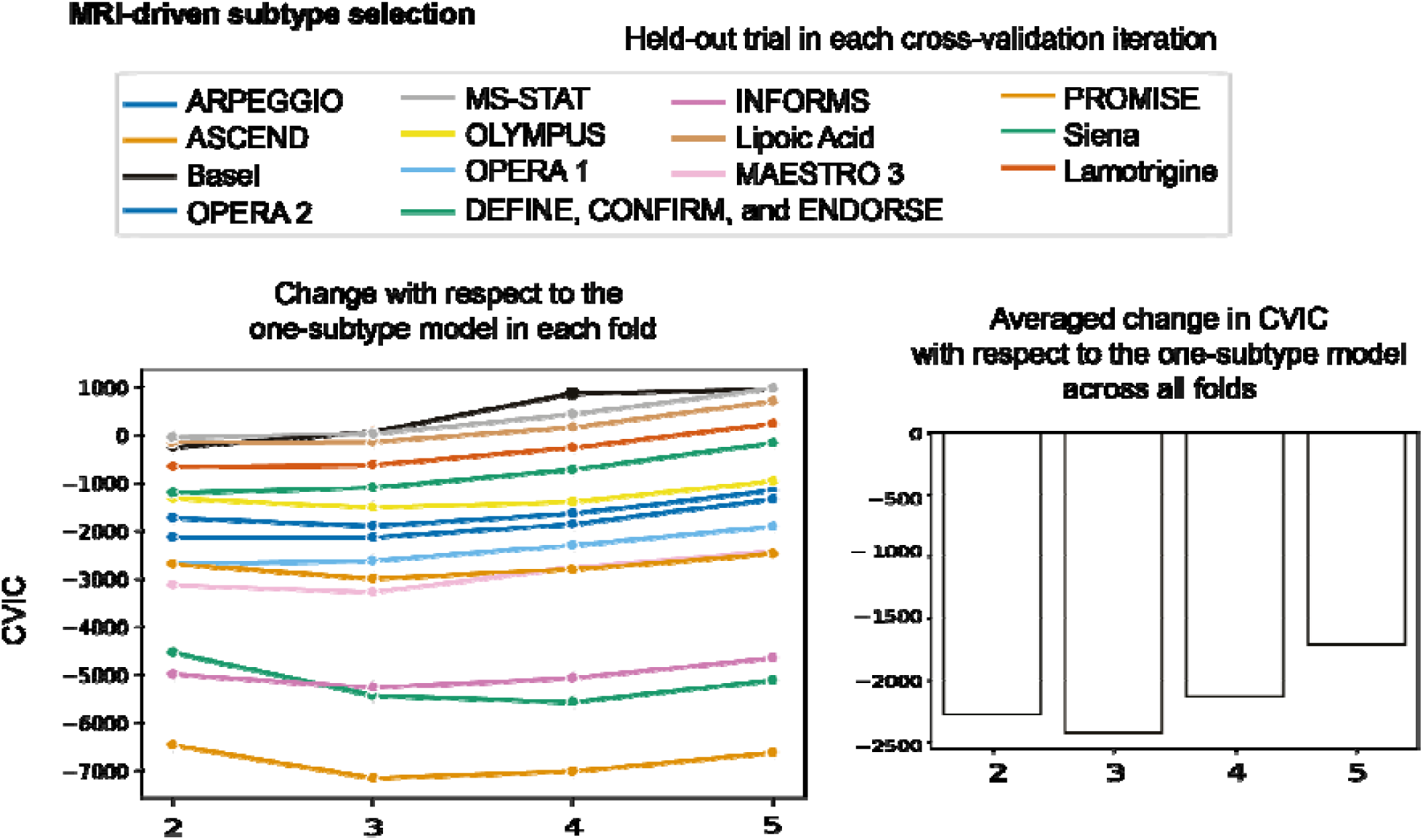
Leave-one-dataset-out cross-validation and model selection. We used leave-one-dataset-out in the train set of 14 studies, each time leaving one study (or dataset) out and fitting SuStaIn algorithm on the remiaing 13 datasets. We chose the best number of subtypes according to the cross-validation information critiera (CVIC) calcualted from the *left-out* dataset each time (x14). The vertical axis shows the change CVIC. Absolute greater increases favour better models: we found that the three-subtype model was the optimal model.

**Supplementary Figure 3.**
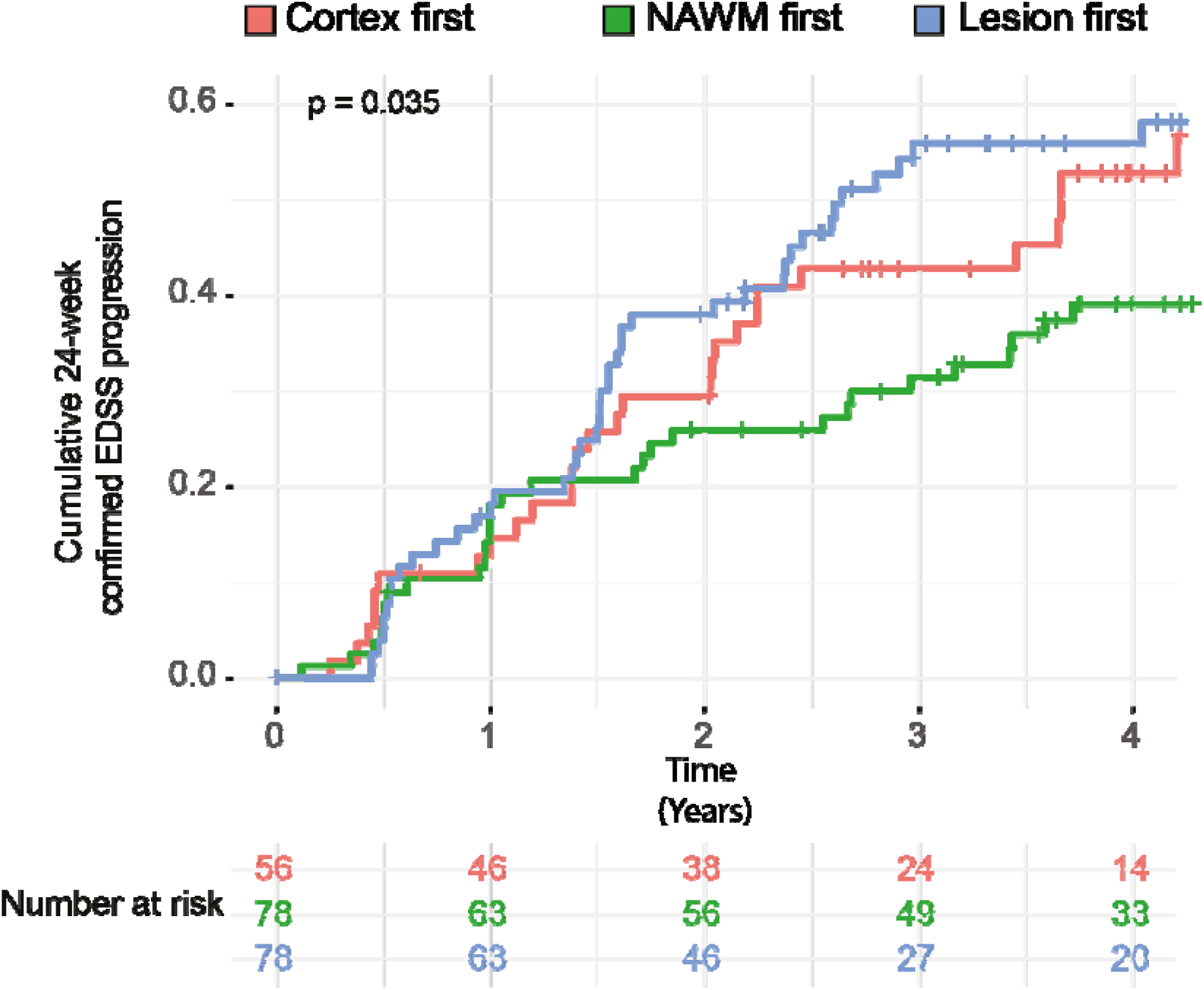
Kaplan-Meier curves showing the proportion of patients reaching 24-week confirmed EDSS progression in each MRI-driven subtype in the CLIMB dataset. *p*-value is for the log-rank test, which compares hazard-ratios of reaching the 24-week confirmed EDSS progression across the three data-driven subtypes. The lesion led subtype had a shorter time to reach this disability milestone compared to the NAWM-led subtype but not the cortex-led subtype.

#### International Progressive MS Alliance (PMSA) Investigators of the network

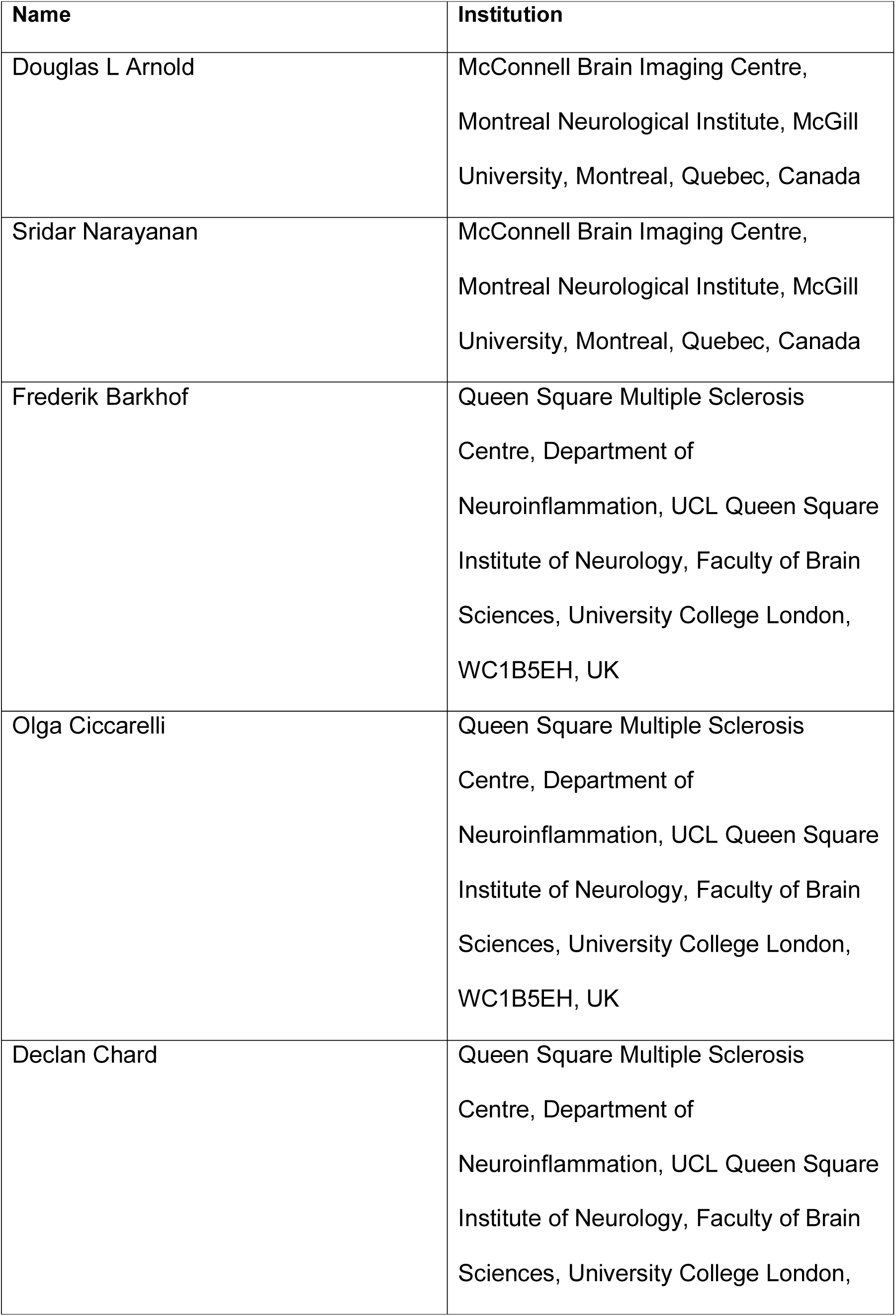
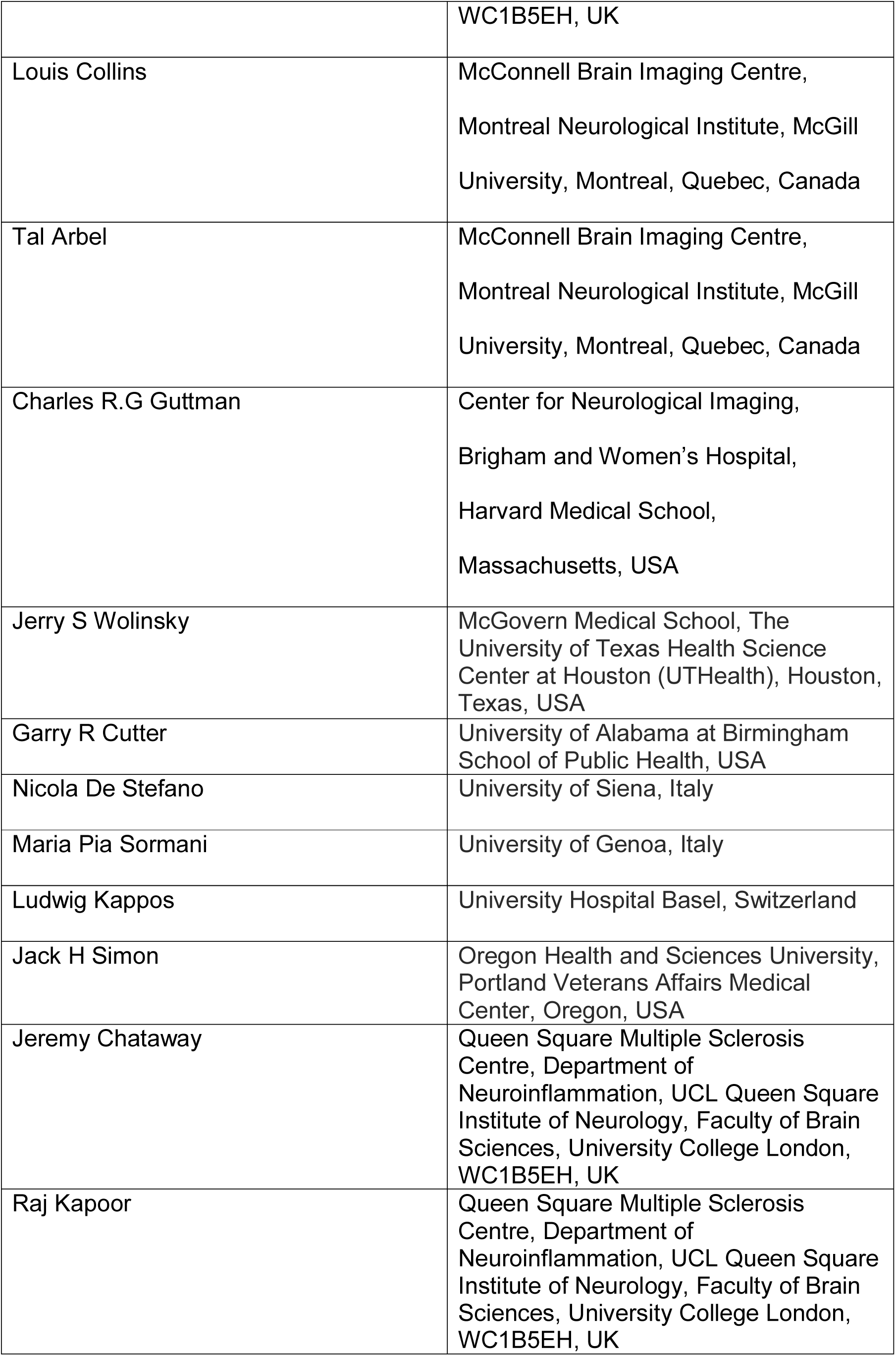
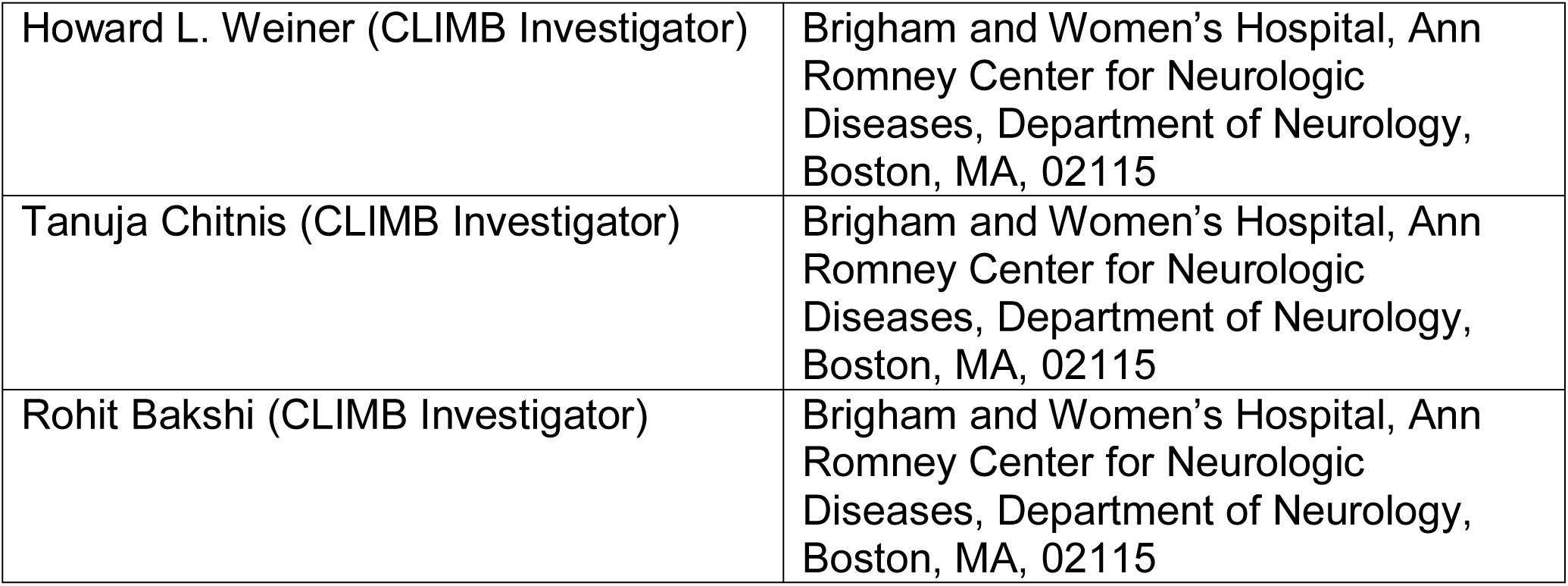

